# Large-scale RF mapping without visual input for neuroprostheses in macaque and human visual cortex

**DOI:** 10.1101/2024.12.22.24319047

**Authors:** Antonio Lozano, Xing Chen, Mike La Grouw, Bingshuo Li, Feng Wang, Maureen van der Grinten, Cristina Soto-Sánchez, Aitor Morales-Gregorio, Eduardo Fernández, Pieter R. Roelfsema

**Author notes:** Correspondence Antonio Lozano & Pieter Roelfsema. Contributed equally (Antonio Lozano, Xing Chen).

## Abstract

High-channel-count neuroprostheses could one day restore functional vision in blind individuals by delivering electrical pulses to electrodes in the visual cortex that elicit perceptions known as ‘phosphenes’. However, if a high number of electrodes are used, it becomes challenging and time-consuming to map the visual field locations of all phosphenes. Furthermore, many blind users are not able to maintain stable fixation, impeding the localization of phosphenes, or may perceive spontaneous visual phenomena that interfere with detection of electrically induced phosphenes. Here, we introduce NEural Unsupervised electrode mapping (NEUmap), a rapid, largely automated method for phosphene mapping that extracts spatial patterns from spontaneous activity across the visual cortex. As correlations between neuronal activity on nearby electrodes are stronger than those between distant electrodes, we first use dimensionality-reduction algorithms to generate maps of relative positions of electrodes. We then convert these maps from relative to absolute visual field coordinates while the subject maps out a small number of phosphenes manually. NEUmap generated maps across ∼300-700 electrodes in each of two sighted monkeys and across 73-91 electrodes in each of three blind human volunteers. We report that the method allows rapid mapping of many electrodes using less than a second of resting-state data, with minimal effort from the subject, in the absence of vision.

## Introduction

Blindness affects ∼40 million individuals worldwide^1^ and results in substantial decreases in the quality of life, including higher unemployment rates and lower income^2–6^. Technological advances in brain-machine interfaces over the past decades in both humans^7–23^ and animals^24–37^ demonstrated that the delivery of electrical currents to the visual cortex via implanted electrodes generates artificial visual percepts known as ‘phosphenes.’ A future visual prosthesis system would comprise a camera on glasses that streams the video feed to a pocket processor for image processing, which in turn sends instructions for electrical stimulation of the tissue. Future users could thereby regain a rudimentary form of vision.

Previous studies showed that phosphene locations closely match receptive field (RF) locations of the neurons in the visual cortex that are stimulated electrically^20,25,27,28,37,38^. Furthermore, a recent study found that monkeys could discriminate more complex phosphene percepts such as letters, which were evoked during the stimulation of combinations of electrodes. Hence, prosthetic vision could potentially convey a recognisable image composed of many simultaneously generated phosphenes. The quality of prosthetic vision should increase with the number of functional electrodes because it increases the number of ‘pixels’ available for the artificially generated images^39–43^. Indeed, a device with hundreds to thousands of electrodes might allow a user to perceive simple shapes and objects in their environment^39,42–54^.

The controlled generation of multiple phosphenes for shape perception requires knowledge of the perceived location of individual phosphenes. In sighted subjects it is possible to map RFs with visually presented stimuli^20,25,27,28,37,38^. However, these RF mapping methods are not possible in blind individuals. In the absence of retinal input, it is possible to estimate the boundaries of visual cortical areas using anatomical methods such as myelin mapping^55–59^, or by examining functional correlations between brain regions based on resting-state BOLD activity^60–62^. In addition, it is possible to coarsely estimate retinotopic maps based on the boundaries between grey and white matter^63–65^. However, none of these estimates yield accurate RF locations of individual electrode sites. Furthermore, there is substantial variability in visual cortical anatomy, e.g. in the size, location, and 3D shape of the visual cortex between individuals^66–72^. Hence, knowledge of the positioning of electrodes in the cortex does not translate into accurate RF predictions^73^.

Instead, researchers have relied on manual mapping methods to determine phosphene locations in blind individuals, for one electrode at a time. The subject reports the location of each phosphene in visual space, e.g. by placing their finger on a touch pad or by drawing its shape and position with a digital pen^8,18,22,23,74–76^. However, these manual mapping techniques have several drawbacks. Firstly, each electrode needs to be stimulated several times to verify the phosphene location via a time-consuming process^8,18,22,23,74,75^ that is unlikely to be feasible for devices with hundreds or thousands of electrodes. Secondly, phosphene locations may be hard or impossible to distinguish when electrodes are near each other^22^. Thirdly, ∼12-59% of blind and visually impaired individuals experience visual hallucinations, which can interfere with the localization of electrically induced phosphenes^22^. Fourthly, many blind individuals exhibit abnormal eye movements^84–86^ that may impair phosphene mapping^87,88^ as the perceived position of a phosphene depends on the direction of gaze^8,10,18,22,89^. These limitations make it unsurprising that the precision of reported phosphene locations is much lower in blind than in sighted subjects^23^.

Here, we describe NEural Unsupervised electrode mapping (NEUmap), a semi-automated phosphene mapping pipeline that exploits distance-dependent correlations in neuronal signals between electrodes (Figure 1). First, resting-state neuronal signals recorded simultaneously from each channel are fed to a dimensionality reduction algorithm (UMAP). This generates a 2D projection, where timeseries data from each channel is assigned a relative position (X, Y) via unsupervised methods, forming a ‘relative map’ of cortical locations across all the channels. Channels with similar patterns of activity will be relatively close together in the relative map. Next, three ‘anchor point’ channels are chosen. Phosphene locations for these channels are manually obtained (in absolute, visual field coordinates), and converted into cortical coordinates. The relative map is then aligned with the cortical projection of the anchors, to yield a map of cortical locations across all the channels. Finally, this cortical map is converted into the visual field coordinates, providing a map of predicted phosphene locations for all channels. We assess the accuracy of the predicted maps by comparing them to ground-truth RF maps obtained in sighted subjects.

**Figure 1.**
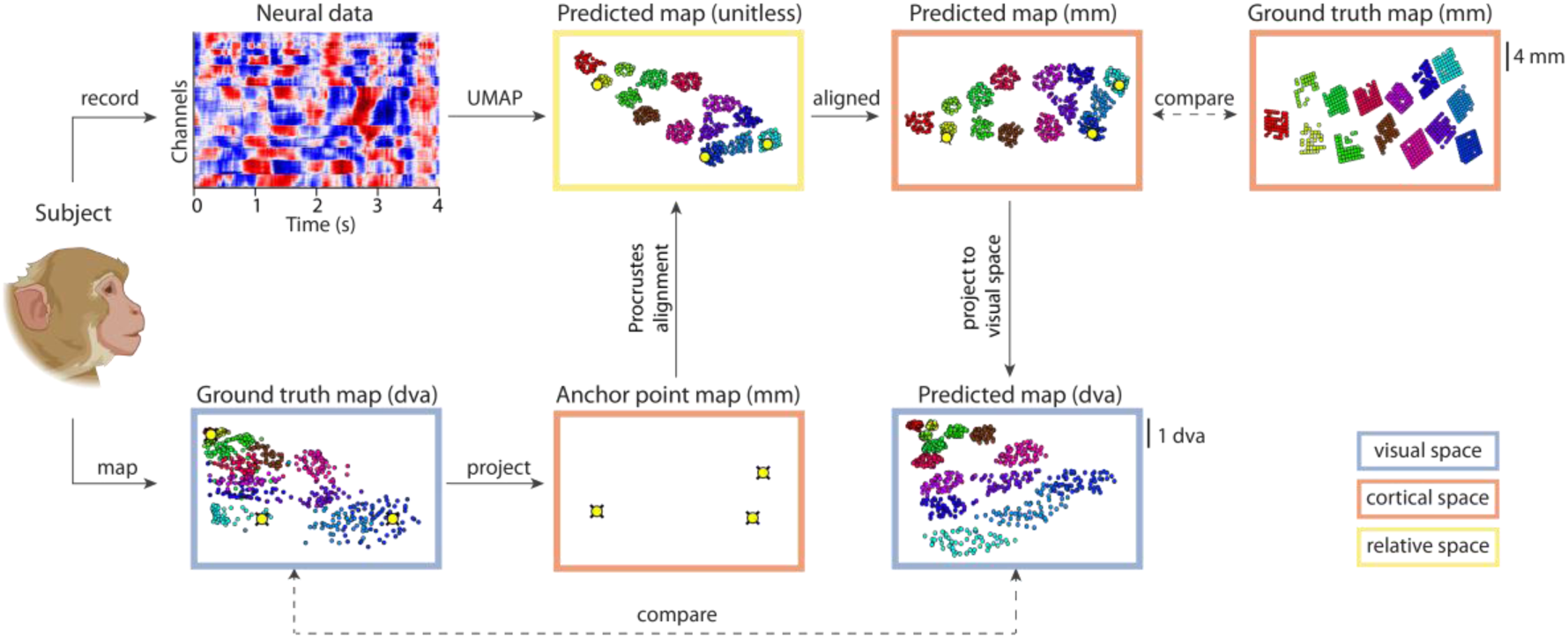
Mapping pipeline. In the monkeys, we recorded neuronal activity (top left) during the resting state and fed these data to UMAP, which assigns a coordinate for each channel within a relative 2D map (‘unitless predicted map’). In the sighted monkeys we measured the RF locations and chose three of them as anchor points (bottom left). The anchor points were converted into cortical coordinates using a wedge-dipole model with parameters obtained from the literature^90,91^. We used the anchor points to align the predicted map in cortical coordinates (‘predicted map’). We compared the predicted map to the ground truth of electrode positions in the cortex determined from pictures taken during the implantation procedure (top right, ‘ground truth map’ in mm). The predicted map was also converted into the visual field coordinates (dva), which was compared to the ground-truth RF map. In blind users, ground truth RF data is not available, and the predicted cortical maps were evaluated against the locations of the electrodes in the cortex. See Supplementary Figure 1 for a detailed view of the differences in available data.

We demonstrated that NEUmap is scalable, successfully yielding RF maps for implants consisting of hundreds of electrodes, with minimal manual input and short amounts of data (∼0.5 s). Thereby, it can decrease the time that prosthesis users spend on phosphene mapping substantially.

## Results

We generated RF maps in the absence of visual input, using our new mapping method, NEUmap. We recorded neuronal data from 14 Utah arrays in the primary visual cortex (area V1) of two monkeys, and a single Utah array in the occipital cortex of three blind human subjects.

We will first present the results obtained in the monkeys, during the resting state with the eyes closed. To this aim we selected epochs when the monkeys were seated in a darkened setup and had closed their eyes^92^. We only included electrodes with a sufficient signal-to-noise ratio (SNR) in the analysis. The SNR quantified the strength of the neuronal response to a checkerboard stimulus. We also excluded electrodes with signs of cross-talk, using a ‘synchrofact participation’ method^92^ (see Methods). In monkey L, the eyes-closed recordings were made at 3 months after implantation, when most of the channels yielded signals with high SNR (*N*=696). In monkey A, the eyes-closed data were collected more than one year after implantation, and fewer usable channels remained (*N*=324). We will also present the results from a larger dataset obtained shortly after implantation in this monkey, while the eyes were open^93^, in a later section.

Figure 2 illustrates the LFP in a time window of 1.5 s. Most power was concentrated in the lower frequency bands (Figure 2E,F)^94–96^. Signals from electrodes of the same array tended to be more similar than those from electrodes of different arrays (Figure 2C, D). We examined how the correlations depended on the distance between pairs of electrodes (monkey L: *N*=241,860 electrode pairs; monkey A: *N*=52,326 pairs, data duration = 180 s). The average correlation decreased as a function of the distance between recording sites, with a space constant *λ* of 5.88 mm in monkey L and 5.89 mm in monkey A (see Methods and Figure 3).

**Figure 2.**
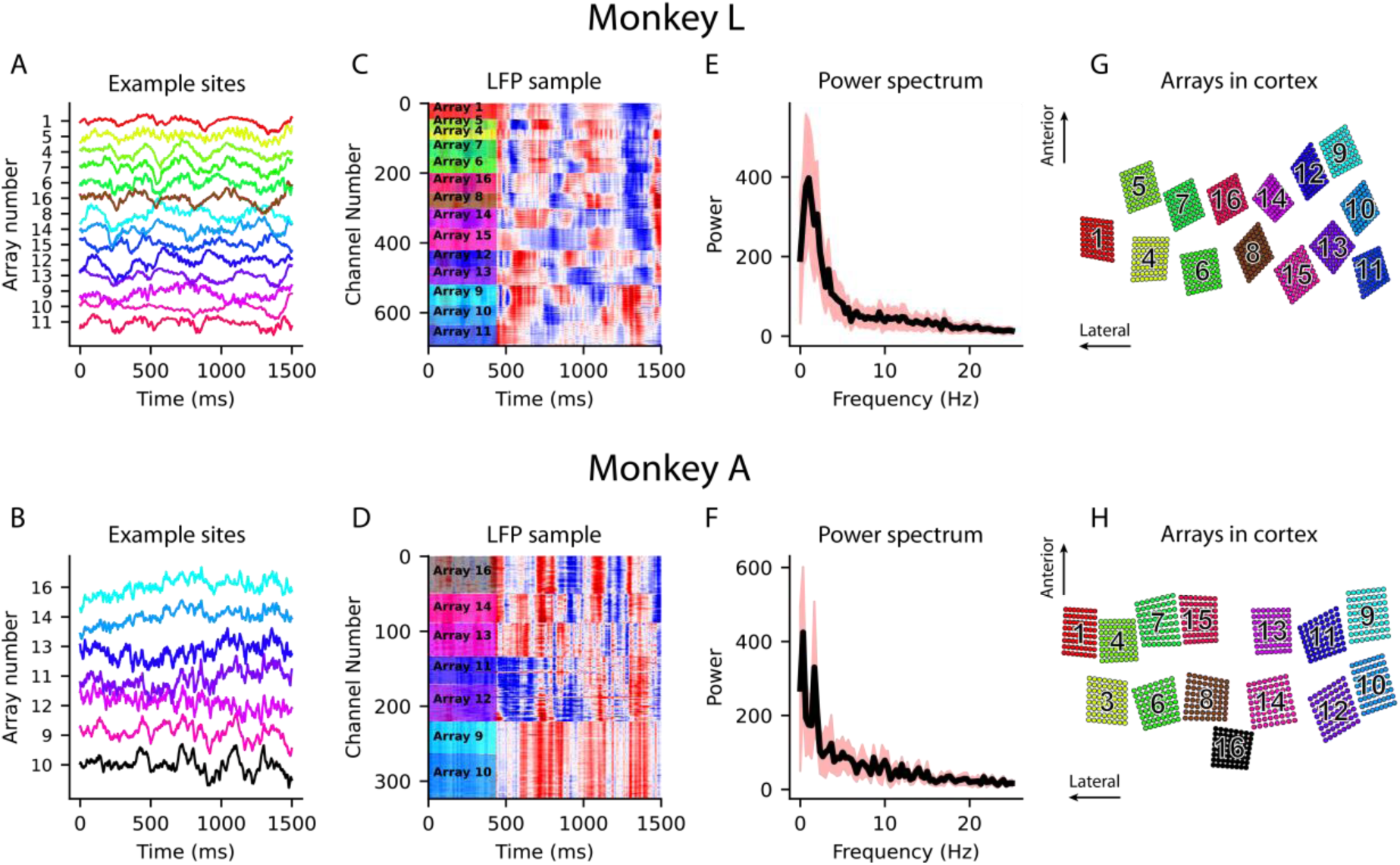
LFP during eye closure. A, B, LFP signals from several example channels in monkey L (A) and monkey A (B). C, D, Broadband LFP data across all included channels (1.5 seconds sample). Positive potentials are shown in red and negative potentials in blue. E, F, Average power spectrum across all included channels. The red shading shows the SD across electrodes. G, H, Positions of arrays in the left visual cortex (V1).

**Figure 3.**
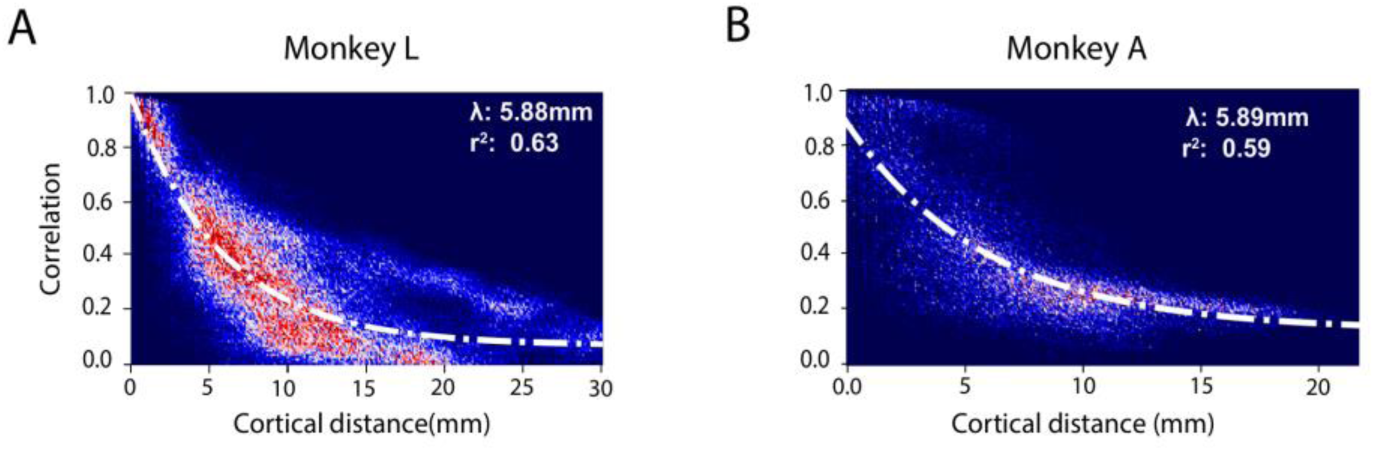
The relationship between the LFP correlation and cortical distance. A, The relation between electrode distance and correlation of the broadband LFP in monkey L (A) and A (B). The fitted curve is an exponential.

We hypothesized that the spatial decay of correlations might differ between the LFP frequency bands and the multiunit activity (MUA) so that they could encode complementary information about interelectrode distances. We measured the strength of signal correlations as a function of distance, for 5 different LFP frequency bands and MUA. The LFP frequency bands were low (1-8 Hz), alpha (8-13 Hz), beta (13-30 Hz), gamma (30-80 Hz), and high gamma (80-150 Hz) (Supplementary Figure 2). For monkey L, the length constant, *λ*, decreased for higher LFP frequencies. For the low and alpha frequencies, *λ* was 5.6 mm; it decreased to 3.3 mm for beta, 3.4 mm for gamma, and 2.4 mm for high gamma, and 1.6 mm for MUA (Figure 4A). In monkey A, *λ* was 4.8 mm for the low LFP frequencies and alpha, 10 mm for beta, 8.4 mm for gamma, 7.7 mm for high gamma, and 6.7 mm for MUA (Figure 4B). Hence, the relationship between the LFP frequency bands and the space constants was not consistent between the monkeys.

**Figure 4.**
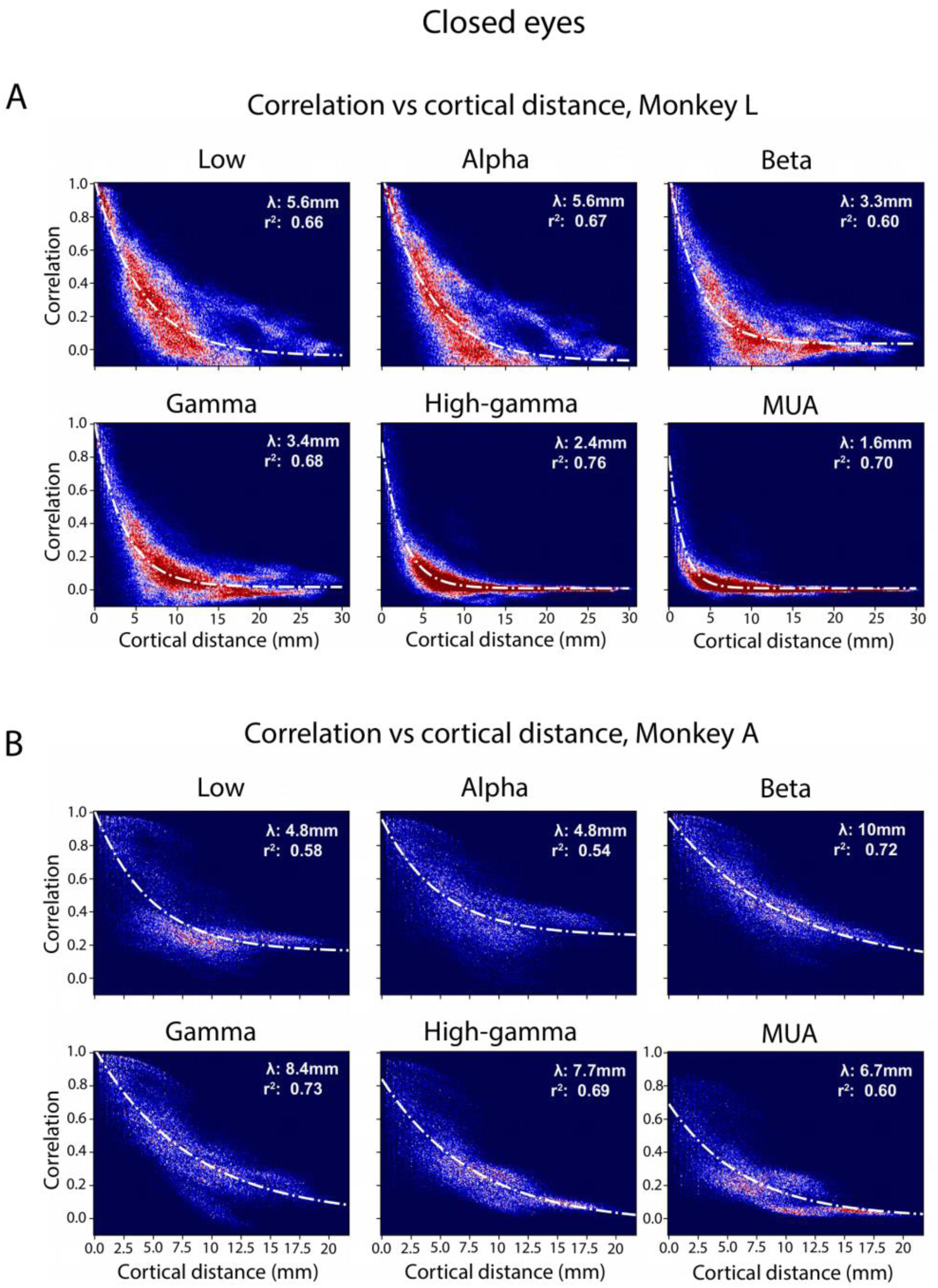
The relationship between electrode distance and signal correlation in 5 LFP frequency bands and MUA, in monkeys L (A) and A (B), using 180 s of data.

The decrease in correlation with cortical distance for both LFP and MUA indicates that these signals could be used to reconstruct the relative positions of the electrodes. We tested both principal component analysis (PCA) and UMAP for dimensionality reduction. We used 15 s of data, sampled at 500 Hz (corresponding to 7,500 samples) so that each electrode contributed a single datapoint in a 7,500-dimensional space. UMAP consistently outperformed PCA (Supplementary Figure 3) and we will report the UMAP results in the rest of the analysis. Supplementary Figures 4 and 5 show the correlation matrices per frequency band in both monkeys.

### Map Predictions with Eyes Closed

We used UMAP^97^, a nonlinear dimensionality reduction method, to estimate the relative electrode locations in a 2D space. We focused on varying two of its main parameters, *min_dist*, the minimum distance between points, and *n_neighbors*, which governs the size of the neighborhood that is considered during optimization steps. The quality of the predicted map depends on these parameters and on the data type, e.g. MUA, LFP or a specific LFP frequency band.

We aimed to select the best UMAP parameters and data type without access to the ground truth, which might not be available in a clinical setting. As a proxy for the ground truth, we therefore used multidimensional scaling (MDS) to create a preliminary map *Map_Prelim_*, based on the pattern of correlations within 15s broadband LFP (see Methods). We chose the UMAP parameters and data type that allowed UMAP to best approximate *Map_Prelim_*.

Figure 5B illustrates the maps based on the gamma-band LFP in both monkeys, which gave the best fit to *Map_Prelim_* (Table 1). Next, we carried out a series of transformations (scaling, rotation, mirroring) to align this map with positions of anchor points in the cortex. We chose three anchor points and used a Procrustes analysis to find the affine transformation that aligned their positions with the cortical coordinates predicted by a wedge-dipole projection^90,98^. We then applied the same affine transformation to all other electrodes in the relative map to obtain an estimated map, *M*_*P*_, across all electrodes in the cortex (Figure 5B) and compared them to the ground-truth locations of the electrodes determined during implantation surgery (Figure 5A). Interestingly, the predicted electrode positions closely matched the ground truth.

**Figure 5.**
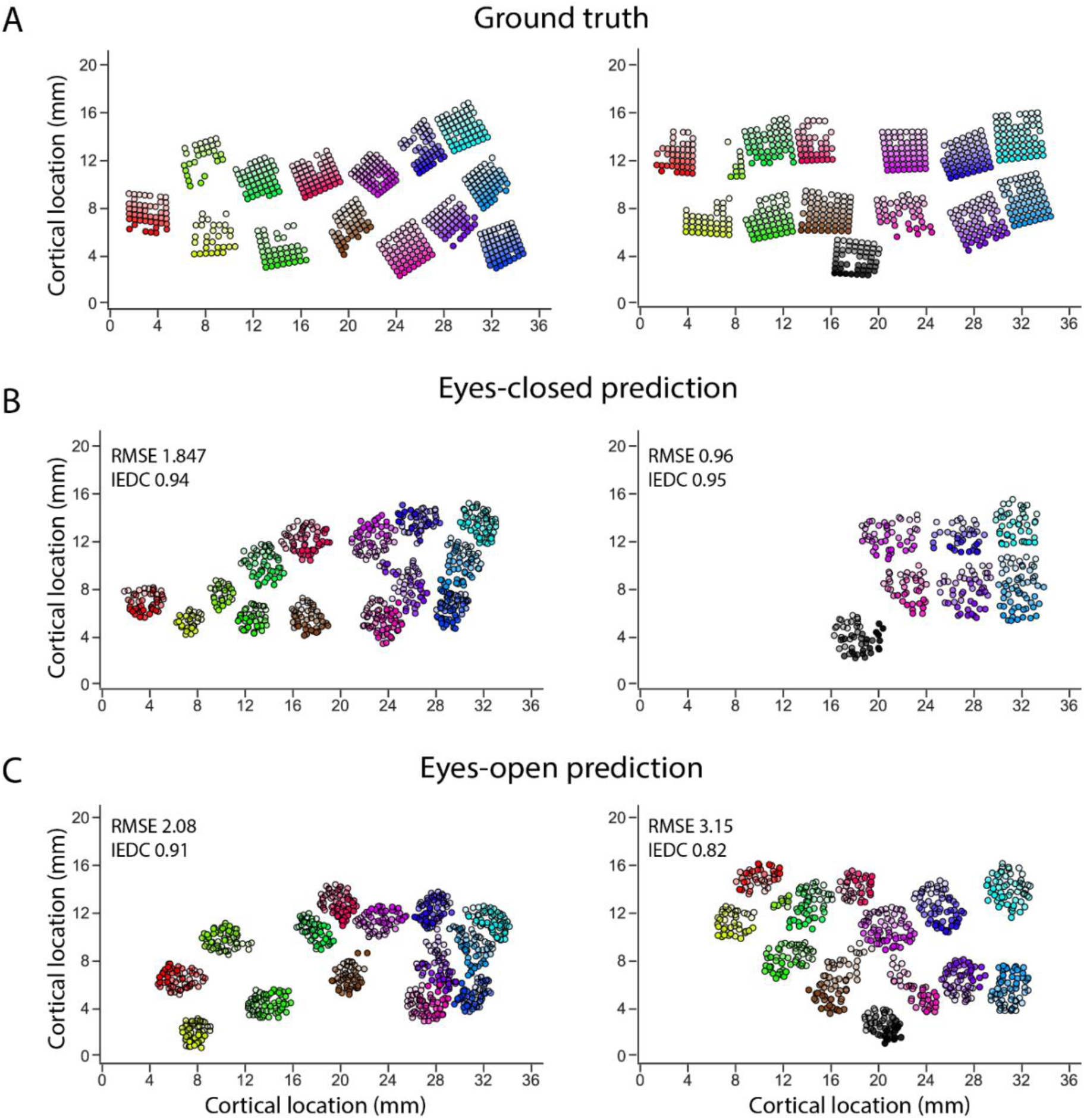
Actual and predicted electrode locations in monkeys. A, Ground-truth cortical locations for monkeys L (left) and A (right), from photographs taken during array implantation. B, Predicted maps when the monkeys had their eyes closed. Based on the fit to Map_Prelim_ we selected the LFP gamma band in both monkeys, and the values of min_dist (Monkey L: 0.5, Monkey: A 0.9) and n_neighbors (Monkey L: 110, Monkey A: 120) (Table 1). C, Predicted maps when the monkeys were looking at a fixation spot on an otherwise blank screen. Here NEUmap selected the beta-band and different UMAP parameters (min_dist both monkeys: 0.9; n_neighbors, Monkey L: 725, Monkey A: 155). Colour saturation indicates array orientation, with lower saturation for electrodes at the more anterior and medial positions. Map accuracy, IEDC, is shown for each example map.

**Table 1:**
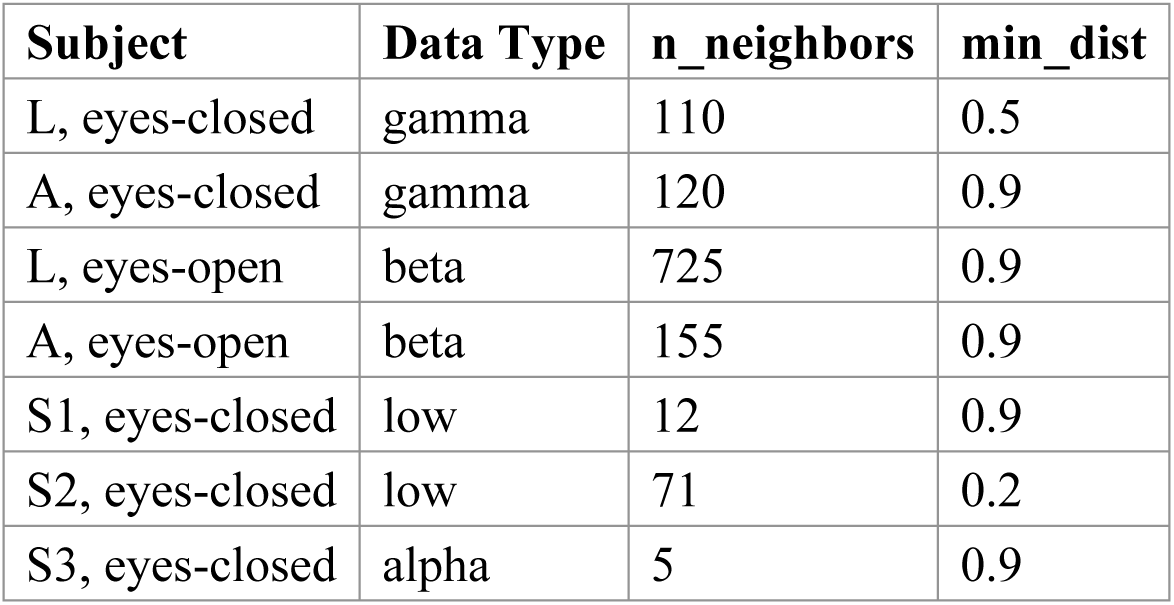
Optimal data types and UMAP parameters for each subject and eyes-closed/open condition.

To test the reliability of the NEUmap, we repeated the procedure 30 times, using randomly selected chunks of 15 s of neural data. To measure the quality of the maps, we computed the inter-electrode distance correlation (IEDC) between estimated pair-wise distances and the ground truth (Supplementary Figure 6). The mean IEDC for gamma LFP was 0.87 ± 0.09 for monkey L and 0.91 ± 0.05 for monkey A. We also measured the accuracy of predicted maps by calculating the root-mean-square error (RMSE) in mm (Methods). The RMSE was 2.8 ± 0.8 mm for monkey L and 1.4 ± 0.4 mm for monkey A (*N*=30 data segments). We also examined the IEDC for the different LFP frequency bands and MUA (Figure 6). For Monkey L, the global map performance was relatively stable across frequencies, but it was lower for MUA when measured within arrays (Figure 6B). In Monkey A, the global performance was high overall, peaking at gamma. The accuracy within arrays was lowest for broadband LFP and MUA (Supplementary Figure 3 illustrates the results of PCA and with the eyes-open data).

**Figure 6.**
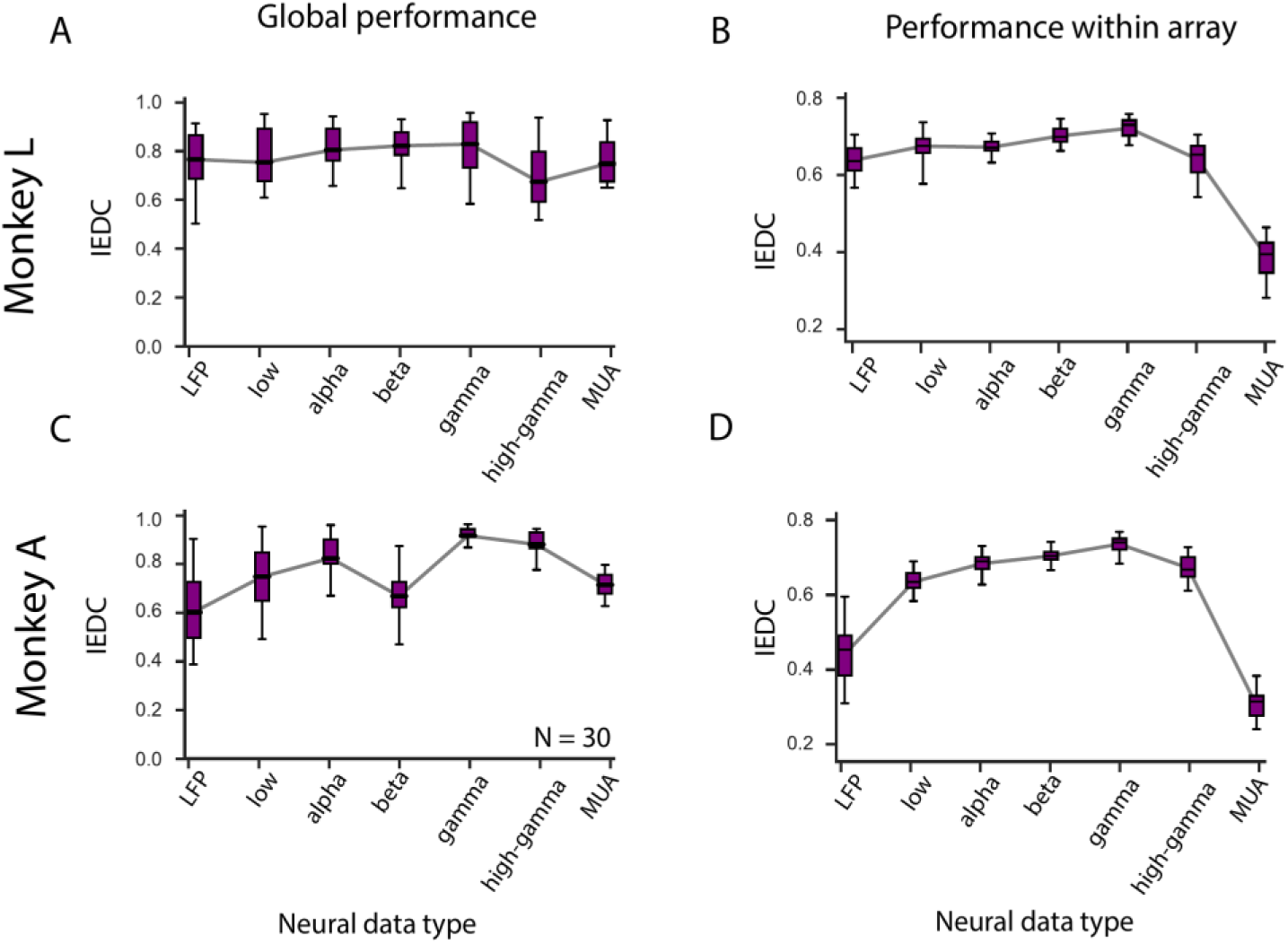
Accuracy of the cortical maps for different frequency bands and MUA when the monkeys had their eyes closed. We measured the IEDC across all arrays (Global performance; A, C) or within individual arrays (B, D), for broadband LFP, 5 LFP frequency bands, and MUA (median and interquartile range).

Thus far, we performed our analyses using data segments with a duration of 15 s. We next examined how the accuracy of the predicted maps depends on the duration of the data segment. We used the best-performing gamma frequency band and UMAP parameters, as was described in the above. NEUmap reached asymptotic performance with as little as 0.5s gamma-band LFP data with an IEDC of 0.9 (Figure 7A,C).

**Figure 7.**
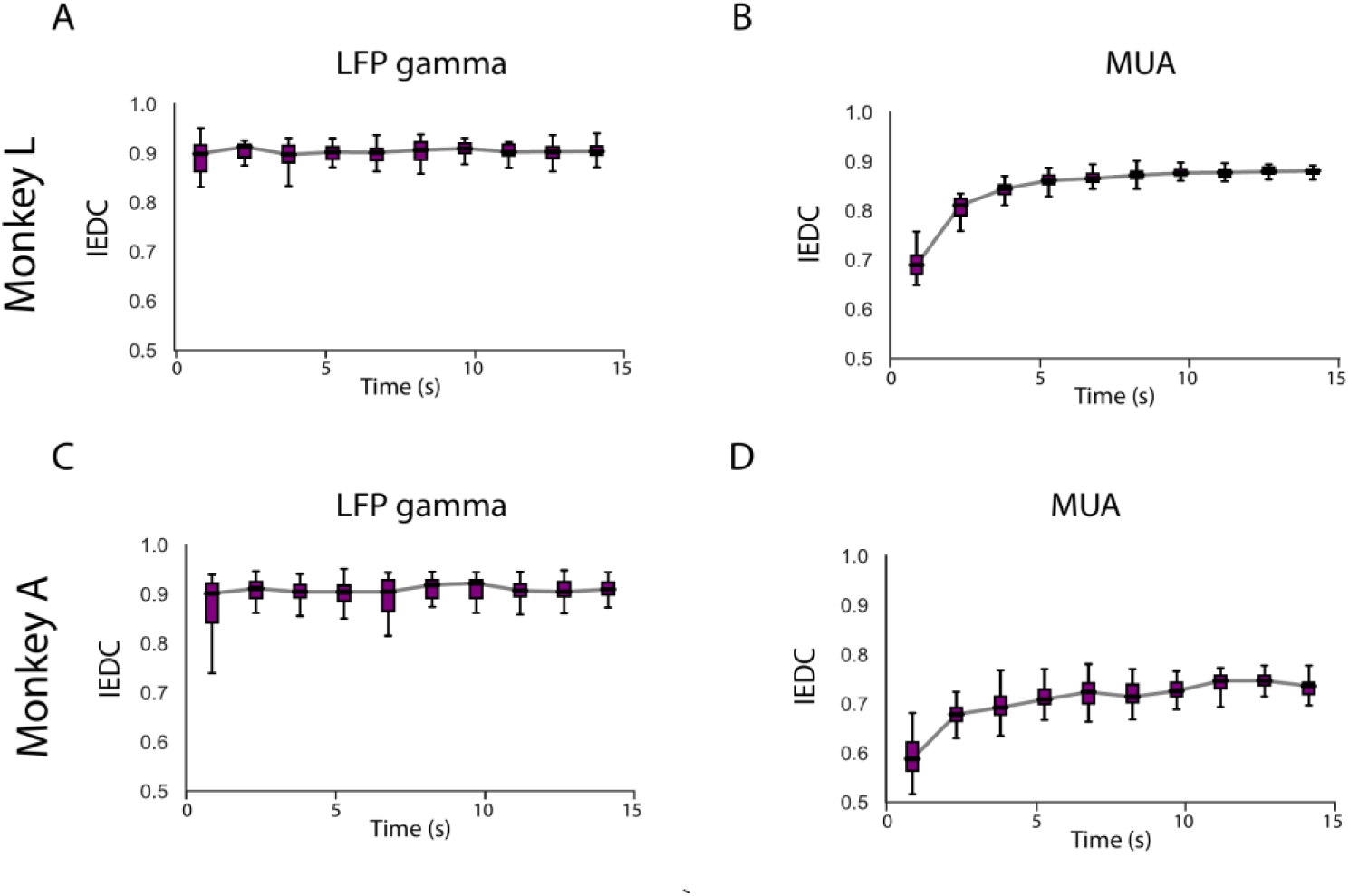
Map accuracy as function of data duration. IEDC for monkeys L (A, B) and A (C, D) using LFP gamma and MUA. The error bars show (median and interquartile range, N = 30 iterations).

We also tested the dependence on the length of the data segment for MUA (Figure 7B,D). The accuracy of the maps increased more gradually for MUA and they reached a stable value after approximately 8s (Figure 7).

### Map Predictions with Eyes Open

The eyes-closed data of monkey A were obtained more than a year after array implantation, when the signal quality of several arrays had degraded^93^, which could explain the lower accuracy of maps based on MUA. We also used NEUmap on data obtained 1-3 months after array implantation, when the SNR was still larger than 2 for 837 electrodes in monkey L and 683 electrodes in monkey A. During these recordings, the monkeys performed a visual task, in which each trial started with a 300-ms fixation epoch during which the monkey directed gaze to a spot at the center of an otherwise blank screen so that the RFs were empty. We obtained 15 s of data by concatenating 300-ms data segments. NEUmap selected the LFP beta-band for these data, based on the fit to *Map_Prelim_* (*N*=30 independent 15-s segments).

As with the eyes-open condition, we observed a decrease in pairwise signal correlations with cortical distance, for both LFP and MUA (Supplementary Figure 7). The predicted maps resembled the ground truth (Figure 5C). Indeed, the IEDC in the eyes-open condition was 0.82 ± 0.08 for monkey L and 0.79 ± 0.09 for monkey A (N=30 repeats). The values were slightly lower than those for the eyes-closed condition (unpaired *t*-test, monkey L: *t*(58) = −1.88, *p* = 0.0647; monkey A: *t*(58) = −6.76, *p* < 0.0001). The RMSEs were 3.5 ± 0.7 mm for monkey L and 3.7 ± 0.7 mm for monkey A, respectively. Again, the RMSE was higher than under eyes-closed conditions (unpaired *t*-test, monkey L: *t*(58) = 2.79, *p* = 0.0071; monkey A: *t*(58) = 15.03, *p* < 0.0001). Hence, NEUmap also generated excellent RF maps across >600 electrodes in both monkeys, although the accuracy was higher when the eyes were closed than when they were open.

### Maps of Single Arrays in Humans and Monkeys

Thus far, we generated RF maps across multiple arrays in the visual cortex of monkeys. Next, we examined the accuracy of maps generated for individual arrays, in the eyes-closed condition. We could carry out this analysis in monkeys and humans. The three blind human volunteers S1, S2 and S3 were implanted with one 96-channel Utah array, and we predicted the locations of 93, 45, and 71 electrodes, respectively (after exclusion of outlier channels, see Methods).

We randomly selected data segments of 15 s and applied NEUmap to data from individual arrays (N=30 repeats). The predicted cortical locations had a grid-like arrangement that matched the actual locations of electrodes within the arrays (Figures 8A,B). The IEDC was 0.89 ± 0.04, 0.93 ± 0.03, and 0.89 ± 0.02 for subjects S1, S2, and S3, respectively (Figure 8C). Similarly, the average IEDC for monkeys L and A were 0.81 ± 0.10 and 0.72 ± 0.12, respectively. Thus, the patterns of spontaneous activity in the visual cortex were informative about the relative locations of electrodes in the sighted monkeys, and also in humans after years of blindness.

**Figure 8.**
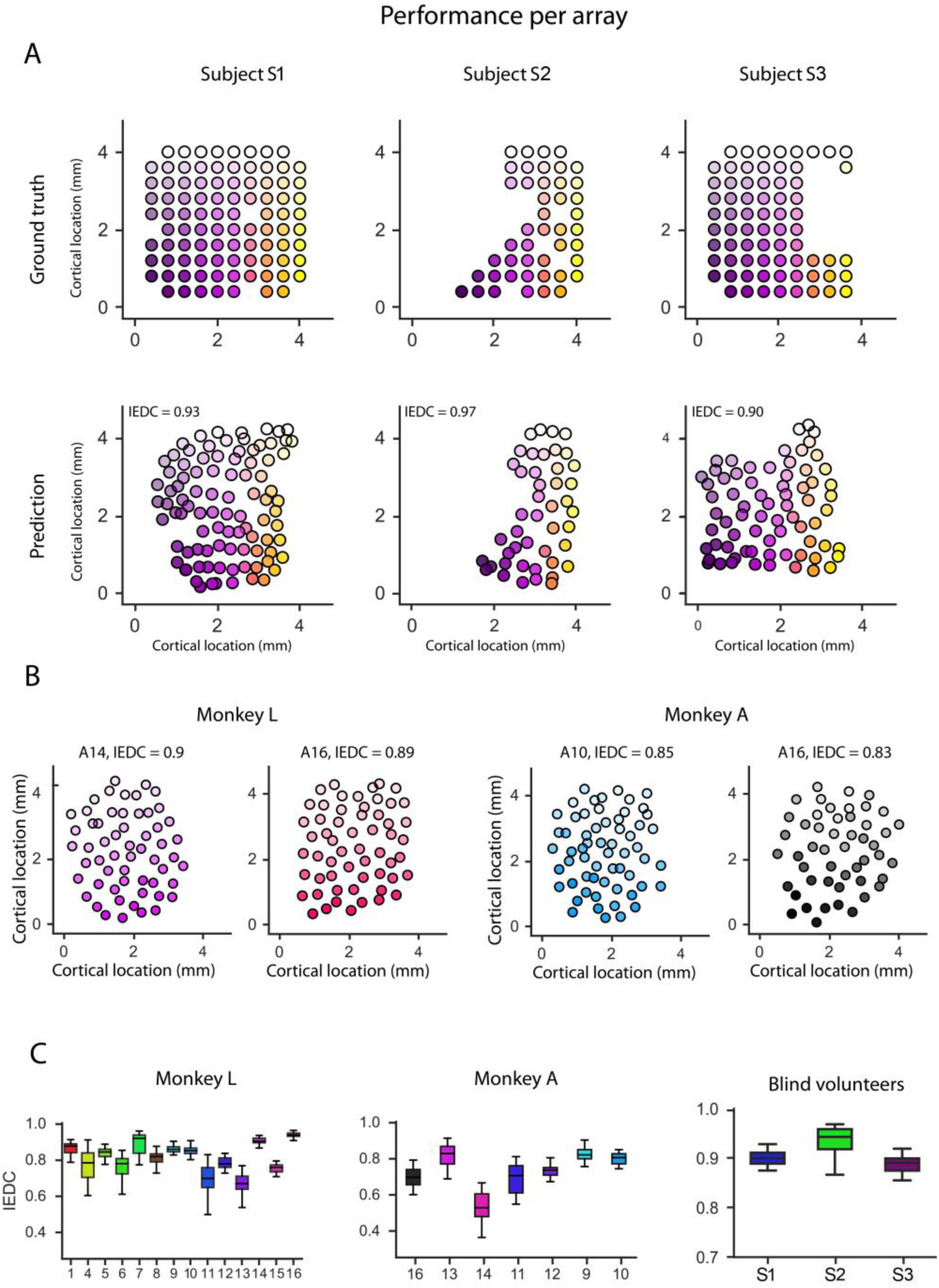
Accuracy of the predictions for individual arrays in monkeys with eyes closed and blind human subjects. A, Ground-truth electrode locations after removal of electrodes without good signal (top) and maps predicted using LFP data in three blind human subjects (S1, low; S2, low; S3, alpha band). B, Predicted maps for example arrays in monkeys. Colour saturation indicates array orientation, with lower saturation for anterior and medial positions. C, IEDC values for the arrays in monkeys L (left) and A (middle), and blind subjects S1, S2 and S3 (right). Note that the Y-axis range differs for the right panel to aid visualization.

### Conversion into Visual Field Coordinates

We applied a wedge-dipole model^90,91^ to the data of the sighted monkeys, to assess the similarity between measured and predicted RF locations. Specifically, we converted the ground-truth cortical maps and the NEUmap prediction (Figure 9A, B, C) into a retinal reference frame (Figure 9 D, E). These maps corresponded well to the RF locations that were measured with visually presented moving bar stimuli (Figure 9F shows the results in Monkey L and Supplementary Figure 8 those in Monkey A).

**Figure 9.**
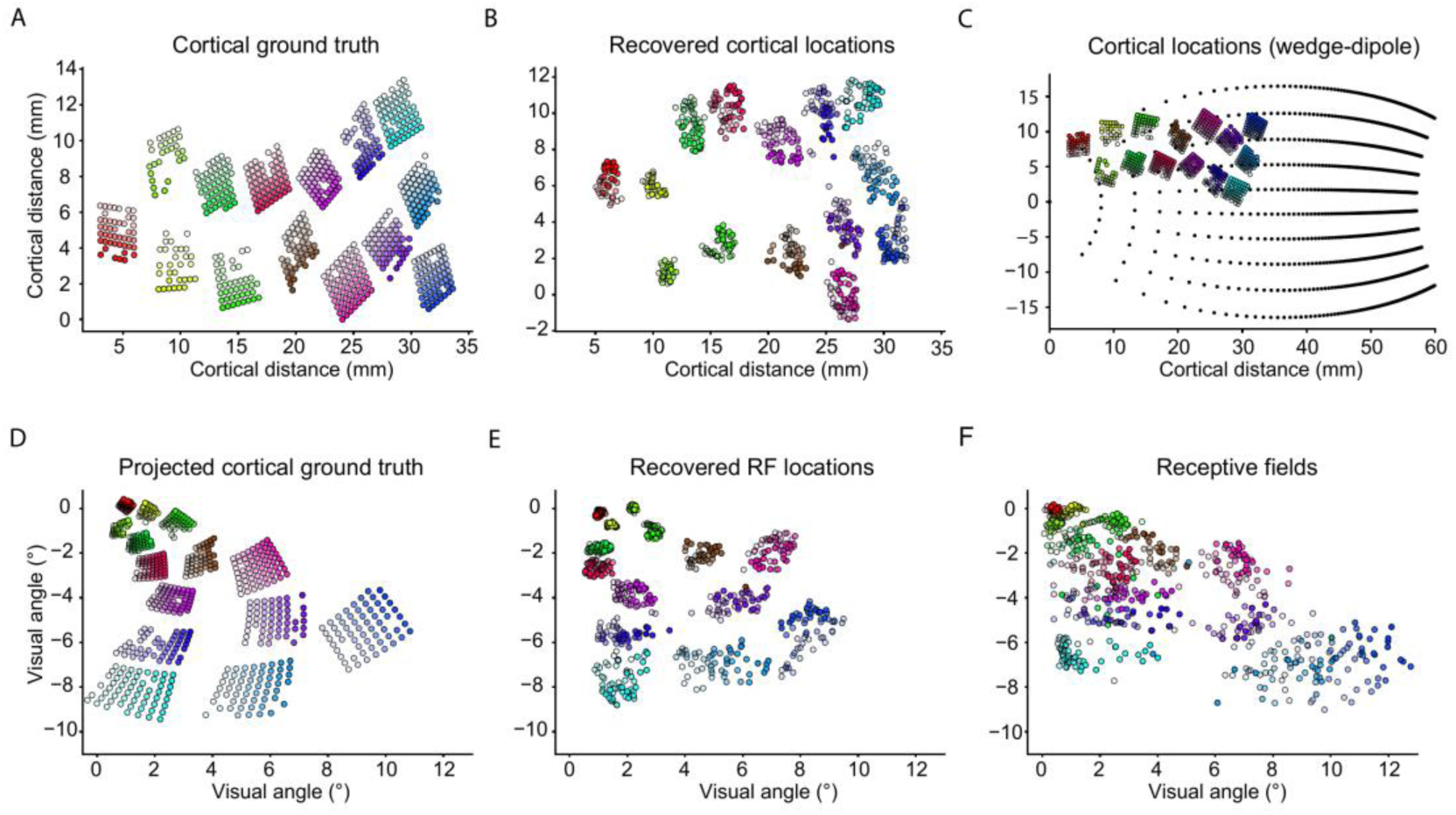
Transformation into visual field coordinates. A, Positions of electrodes in the cortex in monkey L, determined during surgery. B, NEUmap prediction in cortical coordinates. C, A wedge-dipole model was used to convert cortical coordinates into retinotopic space. D, Visual field coordinates obtained by conversion of the array locations. E, Visual field coordinates obtained by the conversion of the cortical coordinates of NEUmap. F, RFs measured with a moving bar stimulus while the monkey looked at a fixation spot.

### Mimicking Discontinuous Cortical Maps

The electrode arrays tiled a continuous region of V1 in the monkeys. However, the three-dimensional shape of the human visual cortex is complex^66,70,71^, with a significant portion of V1 located along the banks and depths of the calcarine sulcus. In future prosthesis applications, implanted arrays may be positioned predominantly on gyri, because their implantation in the sulcus may be impractical. This approach would cause interruptions in the cortical map. To investigate the effects of such discontinuities on the accuracy of NEUmap’s predictions, we examined the effect of systematically excluding electrodes on both sides of an anterio-posterior axis drawn through the middle of the implantation site in monkey L. We tested exclusion zones of up to 22 mm (Figure 10A), systematically reducing the number of mapped electrodes from 696 to 241.

**Figure 10.**
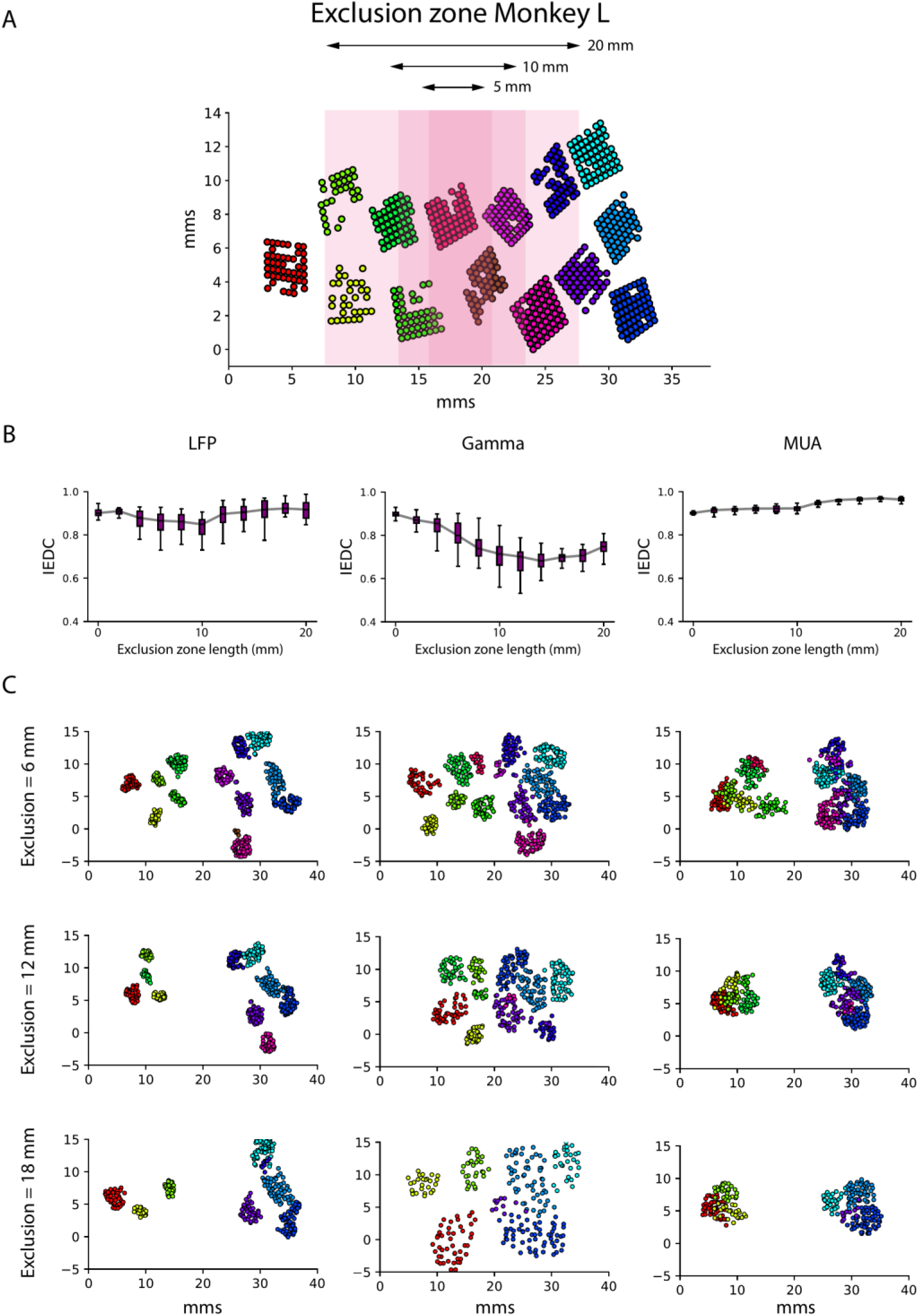
Map prediction accuracy as a function of gap size (median and interquartile range). A, We excluded data from electrodes in a cortical region that varied in size (up to 22 mm). B, EIDC for broadband LFP, gamma-band LFP and MUA as a function of the size of the exclusion zone. Error bars represent SD across 30 data segments. C, Example maps for varying widths of the exclusion zone for broadband LFP, gamma-band LFP, and MUA data.

The quality of the reconstruction depended on the type of data used. For gamma-band LFP, the mean IEDC decreased with the width of the exclusion zone and the positions of the non-excluded electrodes tended to invade into the exclusion zone (Figure 10B). By contrast, the mean IEDC remained relatively stable for broadband LFP and MUA, regardless of the gap size (Figure 10B). These results suggest that the structure of the correlations in broadband LFP and MUA data allows accurate map recovery even when the coverage of the cortical map is discontinuous.

## Discussion

An accurate map of phosphene locations is essential for generating useful perceptions with a cortical implant. Previous work in humans has demonstrated that fMRI can provide coarse maps in blind individuals^60,99^. However, these maps are presumably too coarse for the accurate activation of electrodes if the aim is to generate the perception of fine patterns.

Mapping the precise position of phosphenes can be achieved following electrode implantation, by asking the user to report their perceived location with a pointing device or on a touch pad. However, this procedure becomes very time-consuming when dealing with hundreds or thousands of electrodes. Furthermore, the eye movements of the subjects^87,88^ complicate the manual mapping of phosphene locations, because the perceived location of the phosphene depends on the eye position. Researchers have therefore proposed to train blind subjects to maintain fixation^88,100^ or to use an eye tracker to compensate for changes in eye position^47,101^. In addition, spontaneous visual hallucinations that occur in a fraction of blind individuals can also complicate phosphene mapping, because they are sometimes mistaken for electrically induced phosphenes. Here, we developed a complementary method, NEUmap, that can estimate the relative cortical locations of many electrodes based on correlations in LFP or MUA signals without the active participation of the user.

We tested NEUmap in two sighted monkeys and compared the predicted visual field maps to RF maps obtained using visual stimuli, and the predicted cortical maps to the array locations in intraoperative photographs. In the monkeys, the electrodes were distributed across approximately 300 mm^2^ of V1, and the method yielded reasonable estimates with as little as 0.5 s of LFP-gamma data or 8 s of MUA data. Interestingly, broadband LFP and MUA were advantageous when simulating the implantation of arrays in discontinuous cortical regions.

The pattern of correlations that is exploited by NEUmap provides information about the relative locations of the electrodes that needs to be anchored in the visual field or in the cortical space. Here we only used only 3 anchor points, which is the minimum necessary for alignment. We note, however, that future studies could improve the quality of the maps by adding more anchor points and these anchor points might also help to alleviate distortions caused by electrode implantations in discontinuous cortical regions. In such cases, NEUmap could be used to predict the locations of the phosphenes that are not mapped manually, saving substantial time compared to mapping all phosphenes manually.

To ensure the clinical validity of our approach, we also tested NEUmap on resting-state data from single Utah arrays that had been implanted in the occipital cortex of 3 blind human volunteers. The predicted electrode locations in the humans closely matched their actual layout on the arrays, just as was the case for the monkeys.

## Future Directions and Limitations

We tested NEUmap on data from electrodes in area V1, but expect that the method is equally applicable to other brain regions. Stimulation of extrastriate visual areas such as V2^16^, V3^16,29^, V4^16^, and MT^29^ also elicits phosphenes, and future visual prostheses may implant electrodes in multiple visual areas to extend the coverage of the visual field and generate richer percepts^28,102^. Future studies could examine whether NEUmap provides accurate maps when electrodes are placed in multiple visual areas or in regions without a clear retinotopy, such as the inferotemporal cortex. It is likely that the method could also be used in other brain areas targeted for sensory restoration and decoding applications, including the (pre)motor^103–105^, somatosensory^106,107^, auditory, and (pre)frontal cortices, as well as sub-cortical brain regions like the LGN^108^.

A previous study with data from same monkey revealed a number of differences in the neuronal activity between the eyes open and eyes closed conditions^96^, including changes in the correlations between areas V1 to V4. In the present study, we found that the mapping procedure worked better with the neuronal activity while the eye were closed. These results suggest the pattern of correlations better reflects the local network connectivity when the eyes are closed.

A limitation of NEUmap is that the accuracy of the predictions depends on the quality of the neuronal signals, and also on the parameters and initialization of the method^109^. We addressed this by carefully selecting channels based on SNR. We used the synchrofact detection to remove data chunks with artifacts to ensure data quality. We selected the best data type and UMAP parameters based on the quality of the approximation to an initial RF map using multidimensional scaling, without accessing the ground truth. Future work could explore additional methods to improve map prediction, through either unsupervised or semi-supervised dimensionality reduction methods^110,111^. Future work could also examine whether the combination of LFP/MUA features, and dimensionality reduction with pre-trained neural foundation models^112^ could improve the quality of the maps. Furthermore, it might be possible to combine NEUmap with methods for the optimization of pre-surgical electrode placement planning^73^.

Furthermore, if brain implants are capable of both electrical stimulation and recording, researchers could also examine the amplitude of the neuronal response elicited by electrical stimulation as a measure of cortical distance^115^.

A reliable map of the position of all phosphenes will be essential for the transformation of visual information into artificial percepts. NEUmap aids in the construction of these maps, by accurately predicting the relative cortical locations of electrodes, and by converting them into visual field coordinates. The method is applicable to individuals who have been blind for decades, reducing the time and effort required for manual phosphene mapping. We hope that NEUmap will facilitate the clinical use of future high-channel-count neuroprostheses so that they can aid in tasks of daily living^116,117^ and assist with the personal needs of blind users^118,119^.

## Methods

We used data from two sighted monkeys and three blind human subjects. In the monkeys, we recorded neuronal activity from 896 electrodes across 14 Utah arrays (Blackrock Neurotech, 64 electrodes per array) located in V1^37,92,120^. The monkeys were either in the resting state with their eyes closed (‘eyes-closed’ condition) or they directed their gaze to a spot in the center of an otherwise blank computer screen (‘eyes-open’ condition). In the three human volunteers, we recorded from 96 electrodes of a Utah array that was positioned close to the V1/V2 border in the visual cortex^22^. The human subjects kept their eyes closed throughout recording. (See Supplementary Figure 1 for an overview of the dataset.)

We extracted two types of neuronal signals: multiunit activity (MUA) and local field potentials (LFP). MUA reflects the spiking of neurons within 100 to 150 *μ*m of the electrode, and MUA population responses are very similar to those obtained by pooling across single units^121–124^. To generate MUA, the signal was filtered between 0.5 and 9 kHz. Full-wave rectification was performed on the filtered signal, followed by low-pass filtering at 200 Hz (Butterworth filter, order 4). The data were down-sampled by a factor of 60, yielding MUA with a 500-Hz sampling rate. To generate LFP, raw data were low-pass filtered at 150 Hz (Butterworth filter, order 4) and down-sampled to 500 Hz.

### Monkey Recordings

All experimental surgical procedures complied with the NIH Guide for the Care and Use of Laboratory Animals (National Institutes of Health, Bethesda, Maryland) and were approved by the Institutional Animal Care and Use Committee of the Royal Netherlands Academy of Arts and Sciences (approval number AVD-8010020171046). Two male rhesus macaque monkeys (Macaca mulatta, monkeys L and A) each received two cranial implants during separate surgical procedures, as previously described^92^.

The first implant was a customized, 3D-printed head post for head fixation^125^. The subjects were 4 and 5 years old, weighing 6.5 and 7.2 kg, respectively, at the time of head post implantation. The second implant was a 1024-channel interface with the visual cortex, consisting of 16 Utah arrays (Blackrock Neurotech) attached to a customized, 3D-printed pedestal via 7-cm-long wire bundles^37^. Both monkeys were 7 years old, weighing 11.0 and 12.6 kg, at the time of array implantation. We made a craniotomy over the left hemisphere, opened the dura, and implanted 14 arrays in V1 and 2 in V4^93^. Each array was an 8-by-8 grid of 64 electrodes with sputtered iridium oxide tips (shank length of 1.5 mm; shank pitch of 400 *μ*m). The cortical locations of the arrays were documented with intraoperative photographs.

Pre-implantation electrode impedances ranged from 6 to 12 k*Ω* (measured by Blackrock Neurotech). Each electrode was connected to a contact pad on the Land Grid Array (LGA) interface of the pedestal. Reference wires were attached to every other array, and each wire served as the reference for two arrays, yielding eight reference wires in total. Each reference wire exited the wire bundle several millimeters before the point where the wire bundle met the array.

The head post was affixed to the primate chair to stabilize the head. Electrophysiological signals from V1 and V4 were recorded from 1024 channels (sampling rate of 30 kHz; Blackrock Neurotech data acquisition system, see Figure 4 of Chen et al.^92^, for a schematic of the setup). We used an infrared eye tracker (TREC ET-49B, version 1.2.8, Thomas Recording GmbH) to sample pupil position and diameter (frame rate of 230 Hz).

We recorded resting-state data 3 and 13 months after array implantation in monkeys L and A, respectively^92^.The monkeys were head-fixed in an experimental room with the lights turned off. The setup was not completely dark due to the presence of LED lights on our recording equipment and a small amount of light coming from under the door, and the room was quiet. The monkeys did not carry out a task and were awake or fell asleep during the recording. They occasionally exhibited signs of sleepiness, and their eyes sometimes stayed closed for minutes at a time.

We recorded the pupil diameter of the right eye and considered the eyes to be closed if the diameter fell below a threshold. For each segment with closed eyes, we excluded the first 4 s of data to avoid contamination from visually evoked input, and excluded data segments of >4 s in length from the analysis. We then concatenated the data segments, yielding a total of 2844 s (47.4 min) and 1300 s (21.7 min) of eyes-closed data in monkeys L and A, respectively. Signals were *z*-scored for each channel to ensure a comparable range across channels.

We assessed the signal quality (SNR) by recording visually evoked responses to checkerboard stimuli (> 30 trials) during a fixation task. The monkey fixated on a dot at the center of a grey screen (background luminance, 14.8 cd/m^2^) for 400 ms. Next, a full-screen checkerboard stimulus was presented for 400 ms, and the monkey maintained fixation to receive a reward. The size of the checkerboard squares was 1 dva and the luminance of the black and white squares was 0 and 92.1 cd/m^2^, respectively. We computed signal-to-noise ratios (SNRs) as the amplitude of the visually driven response divided by the standard deviation of activity in a time window before stimulus onset^92^ and excluded electrodes with SNR < 2 and/or signs of cross-talk, using a ‘synchrofact participation’ method^92^. In monkey L, 696 channels were included for further analyses (Supplementary Table 1). In monkey A the data were obtained 1 year after array implantation, and 8 arrays had to be excluded due to low SNR, resulting in 324 remaining channels (Supplementary Table 1).

One type of artifact were periods lasting 1 to 10 s in which the signals across multiple channels simultaneously either exhibited high-amplitude artifacts or decreased in amplitude. We detected these periods by *z*-scoring local field potential (LFP) signals within non-overlapping three-s windows. We identified windows where *z*-scored LFP signals 1) were higher than 2 or lower than -2 (‘high-amplitude’ artifacts) for ≥ 0.5 s, or 2) had a standard deviation (SD) of < 0.1 or < 0.2 in monkeys L and A, respectively (‘weak-signal’ artifacts). We excluded time windows in which more than 10 channels had high-amplitude artifacts or more than 20 channels showed weak signals. These exclusion criteria led to the removal of 18% of the data in monkey L and 15% in monkey A.

We also obtained a dataset in which the animals’ eyes were open, at 2-3 and 1-2 months after array implantation in monkeys L and A, respectively. On each trial, the monkeys fixated on a dot located at the center of an otherwise blank screen, prior to the onset of a visual stimulus^92^. We used spontaneous neuronal activity recorded during the 300-ms pre-stimulus epoch, concatenating the 300-ms segments across trials. A total of 837 and 683 channels were included in this eyes-open dataset for monkeys L and A, respectively.

### Research with Human Participants

The study protocol with three human participants was approved by the Clinical Research Committee at the Hospital General Universitario de Elche (ClinicalTrials.gov registration number NCT02983370, ‘CORTIVIS’). All ethical guidelines conformed with clinical trial regulations (EU No. 536/2014 [repealing Directive 2001/20/EC], the Declaration of Helsinki, and the EU Commission Directives 2005/28/EC and 2003/94/EC), and we obtained written informed consent from the subjects prior to the study.

Subject S1 was female in the 55-59 age range who had no light perception for approximately 15 years, Subject S2 was a male in the 60-64 age range with no light perception for approximately 3 years, and Subject S3 was a male in the 60-64 age range with no light perception for approximately 15 years.

A pre-operative 3D model of the occipital cortex and neurovascular structures was constructed using anatomical images acquired on a 3T MRI scanner (Siemens MAGNETON Skyra) using the MPRAGE protocol (192 sagittal slices, 256×256 matrix, 1×1×1 mm^3^, TR = 1900, TE = 2.49, FS = 3). We estimated the retinotopic maps of the occipital cortex based on cortical surface anatomy^64,126^ and predicted the boundaries between visual areas V1, V2, and V3.

A 96-channel Utah electrode array was implanted in each subject’s right occipital cortex (near the occipital pole), close to the V1/V2 border, following standard neurosurgical procedures as described previously^22,48^. The array was a 10-by-10 grid of 96 electrodes with sputtered iridium oxide tips (shank length of 1.5 mm; shank pitch of 400 *μ*m). The final array location was documented with intraoperative photographs, and a postoperative CT scan of its location was merged with the pre-operative model of the occipital cortex.

Neural signals were recorded using a NeuroPort data acquisition system (Blackrock Neurotech) in Subject S1, and the Explorer Summit data acquisition system (Ripple Neuromed) in Subjects S2 and S3. We typically recorded neuronal activity for up to 4 hours per session, with 1-2 sessions per day, for 5 days per week, over a period of 6 months. The subjects were seated in a comfortable chair with their eyes closed. We included data from a single session at 20, 7, and 2 weeks after implantation in Subjects S1, S2, and S3, respectively. The signal durations were 2, 2, and 7.5 minutes, respectively. For Subject S1, we manually excluded three channels by generating an LFP correlation matrix and identifying outlier correlation rows by visual inspection. For Subjects S2 and S3, we identified outlying channels by performing dimensionality reduction with PCA on *z*-scored LFP signals, followed by clustering using HDBSCAN^127^. A total of 93, 45, and 71 channels remained for Subjects S1, S2, and S3, respectively.

### Ground-truth Datasets

To evaluate the performance of NEUmap, we compared the predicted maps to the known cortical electrode locations. In the blind human subjects, we used the grid positions of electrodes on the 96-channel array as the ground truth map. In the monkeys, we 1) mapped the RFs with a moving light bar, and 2) analyzed intraoperative photographs of the arrays on the cortex^37^ (Supplementary Figure 6). We used a wedge-dipole model (described in the following section) to convert phosphene and RF locations from visual field coordinates into cortical coordinates (Figure 1).

To identify channels with good electrophysiological signals, we calculated the signal-to-noise ratio (SNR) of multi-unit activity (MUA) elicited by checkboard stimuli as:

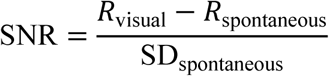

where *R*_visual_ and *R*_spontaneous_ are the responses obtained during and prior to stimulus presentation, respectively, and SD_spontaneous_ is the standard deviation of activity prior to stimulus onset, calculated across trials. Channels with an SNR > 2 were included.

### Transformation from Cortical to Visual Field Coordinates

We converted visual field (retinal) coordinates into cortical coordinates with the wedge-dipole model^90,91^. The location of a point in the visual field can be expressed in polar coordinates *z* = *re*^*iθ*^, where *r* is the eccentricity of the point and (one hemifield). This point is then mapped using the formula:

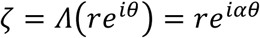

where *α* controls the amount of shear, *∧* is the shearing operation. The wedge-dipole mapping transforms each point to a point *w* in cortical coordinates, using the formula:

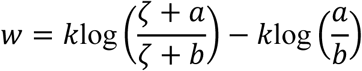

where *a*, *b*, and *k* are parameters determining the two locations of the dipoles and the global scaling of the map, respectively. The second term in this equation ensures that the fovea is located at the (0,0) point in the cortical map. For transformations in the opposite direction, from cortical to visual field coordinates, these mappings are inverted, yielding

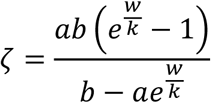

and

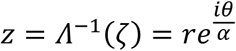

We selected the wedge-dipole map parameters using a grid search to maximize the correlation between photo-derived cortical coordinates and RF-mapping-derived retinal coordinates. Specifically, we calculated the mean correlation between the photo-derived and RF-mapping-derived retinal x- and y-coordinates (x-coordinates: *r* = 0.91; y-coordinates: *r* = 0.88). The best fitting values for parameters *a*, *b*, *k*, and *α* in equation (1) were 0.61, 106, 13.6, and 0.86, respectively.

### Space Constant

To characterize the relationship between correlation and electrode distance, we computed the space constant with the following expression:

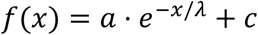

where *x* is cortical distance in mm and *λ* is the space constant in mm.

### Map Computation and Alignment

We used UMAP^97,128^ (Uniform Manifold Approximation and Projection for Dimension Reduction), an unsupervised dimensionality reduction algorithm, to compute a two-dimensional map of the relative position of all recording sites, **M**_*R*_, in relative (unitless) cortical coordinates. To transform **M**_*R*_ into a map with units of cortical distance in mm, we identified the coordinates of a few anchor-point electrodes. Three anchor points suffice for alignment, and we therefore selected three electrodes with receptive fields (RFs) that were spaced far apart (Figure 1). The x- and y-RF coordinates of these anchor points were stored in a matrix **M**_*A*_, and their cortical coordinates as **M**_*R*_*AP*__. We used a Procrustes analysis^129^ to align **M**_*R*_*AP*__ and **M**_*A*_, yielding a scaling parameter *s* and a rotation matrix **R** that minimized the sum of squared point-wise differences between **M**_*R*_*AP*__ and **M**_*A*_. To align the relative map of all the electrodes to **M**_*A*_, we applied the same values of *s* and **R** to **M**_*R*_, obtaining our predicted map **M**_*P*_ in cortical coordinates:

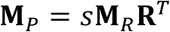

To assess the accuracy of the cortical maps across multiple arrays in monkeys, we used a Procrustes analysis to align the relative map to the ground-truth cortical locations and calculated performance metrics (as described in the next section). To assess the accuracy of maps of individual arrays in monkeys or humans, we used a Procrustes analysis to align the relative map to the physical layout of the Utah array.

### Performance Measures

We evaluated the accuracy of our predicted map, **M**_**P**_, by comparing it to cortical coordinates derived from a photo of arrays on the cortex, GroundTruth_cortical_; as well as to visual field coordinates derived from RF mapping, GroundTruth_visual field_. We calculated the inter-electrode distance correlation (IEDC) between estimated and ground-truth pair-wise distances:

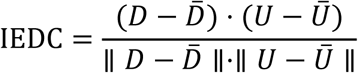

where *D* and *U* are vectors with flattened 0.5 ⋅ *N* ⋅ (*N* − 1) distance estimates and ground-truth distances, respectively, and *D̅* and *U̅* are means. To estimate the error in mm, we also calculated the root-mean-square error (RMSE) between predicted and ground-truth maps as:

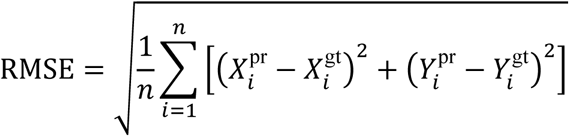

where *n* is the number of electrode locations in each map, *X_i_*^gt^ and *Y_i_*^gt^ are the coordinates of a point in the ground truth map, and *X*_*i*_^pr^ and *Y*_*i*_^pr^ are those in the predicted map.

### UMAP Parameter and Data Type Selection

UMAP^97,128^ is a powerful nonlinear dimensionality reduction method, but it is not guaranteed to recover the global structure of the data^109^. The outcomes depend on two parameters. The first is *min_dist*, which is the minimum distance between points and the second is *n*_*neighbors*, which governs the size of the local neighborhood in UMAP.

We aimed to obtain optimal values for *min_dist* and *n*_*neighbors* without the use of the ground truth data. Instead, we created a preliminary map *Map_Prelim_* based on the correlation matrix that we obtained with 15 s of LFP broadband data and Multidimensional Scaling (MDS)^130^, using the ‘scikit-learn’^131^ implementation. We used *Map_Prelim_* to evaluate the quality of UMAP under various parameter combinations (*n_neighbors*, *min_dist*) using a grid search. We chose *n*_*neighbors* ranging from 10 to *C* − 10, where *C* represents the total number of channels in the dataset and *min_dist* was chosen from [0.1,0.2,0.3,0.4,0.5,0.6,0.7,0.8,0.9]. We initialized the UMAP algorithm with a map obtained by applying PCA to the neural data^109^.

We measured the IEDC between the UMAP result and *Map_Prelim_* 5 times for each parameter combination, and for each neural data type (LFP broadband, LFP frequency-isolated bands, and MUA), subject, and condition (eyes-open and closed). To validate this method to select the UMAP parameters and the best data-type, we calculated the correlation between the RMSE as well as the IEDC between UMAP prediction and *Map_Prelim_*, and that between UMAP predictions and the ground truth. The two measures of error correlated well (range of the correlation coefficient 0.35-0.87; Supplementary Figure 9), demonstrating that *Map_Prelim_* provided a satisfactory estimate during the selection of UMAP parameters and the data type. Table 1 summarizes the selected UMAP parameters and data types, for each subject and condition.

## Data Availability

Data regarding the NHP recordings is publicly available

https://doi.org/10.1038/s41597-022-01180-1

## Acknowledgements

We thank Kor Brandsma and Anneke Ditewig for technical support, Jaap de Ruyter van Steveninck for assistance with wedge-dipole map analyses, and Matthew Self for assistance during surgeries.

## Author contributions

AL, XC, and PR wrote the manuscript with feedback from FW, EF, CS, AM and MG.

PR, XC, and BL supervised and coordinated the project.

Animal research: PR and XC designed the study. XC set up and ran the experiments, collected the data, and generated NHP metadata. AL, XC, ML, MG, AM and BL wrote the data processing scripts, set up the data processing workflow, and managed the data repository.

Clinical research: EF, CS, and AL designed the study. AL and CS created the experimental setup, ran the experiments, collected the data, and generated metadata. AL and MG wrote the data processing scripts, and AL set up the data processing workflow and data repository.

## Supplementary information

**Supplementary Figure 1.**
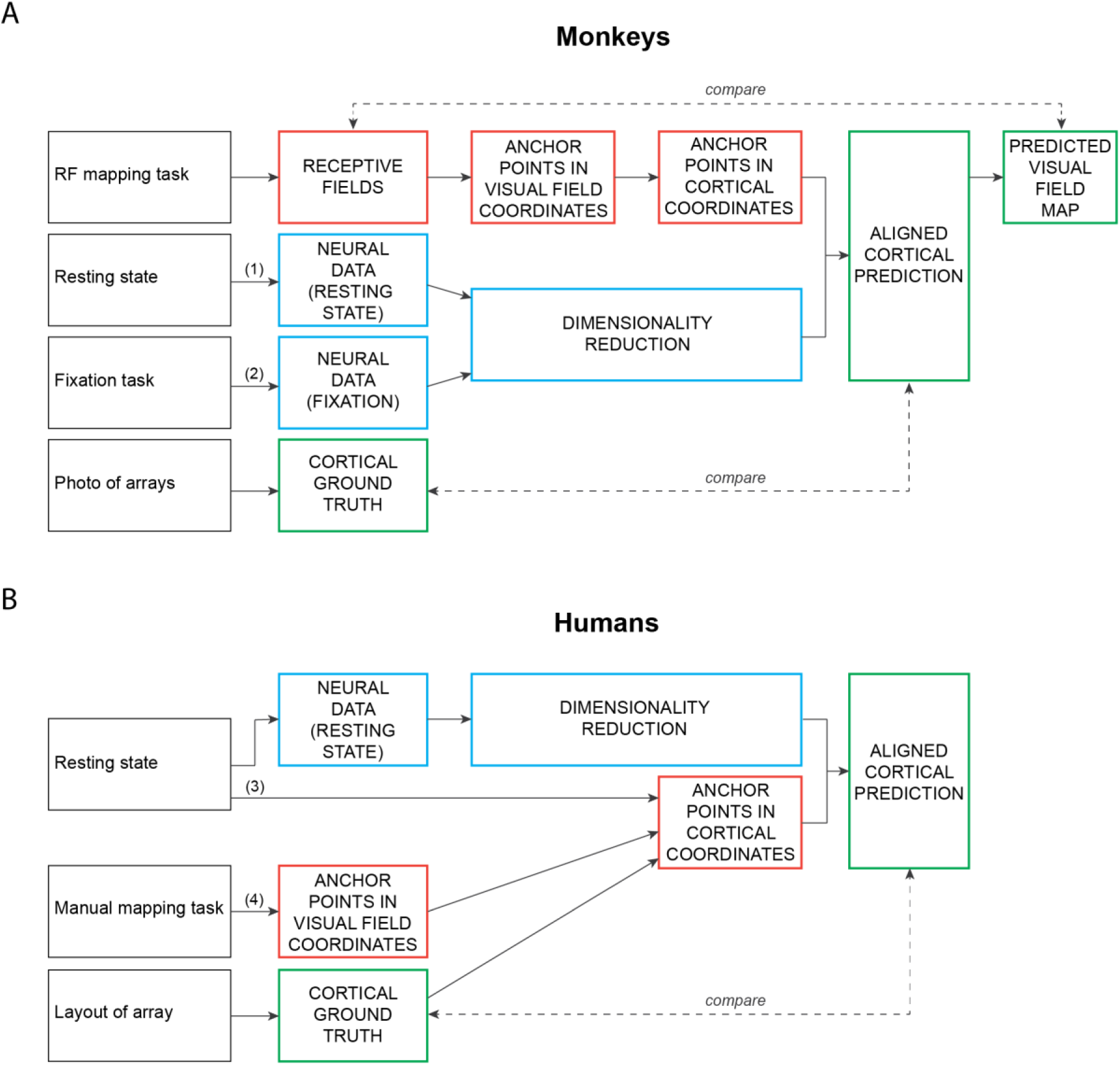
Data collection and mapping pipeline. A, We obtained the following data in monkeys (top): RF maps during an RF mapping task; resting state eyes-closed data; eyes-open data during a fixation task; and an inter-operative photo showing the locations of arrays on the visual cortex. We used UMAP on the eyes-closed and eyes-open data, yielding relative maps, and aligned them to anchor points in cortical coordinates (‘aligned cortical prediction’). The predicted cortical maps were compared to the locations of arrays in the photo; they were also converted into visual field coordinates and compared to the RF maps. B, We obtained the following data in blind patients (bottom): resting state eyes-closed data; phosphene locations during a manual mapping task; and the layout of channel locations on each 96-channel array. We used UMAP and aligned to anchor point coordinates, yielding the ‘aligned cortical prediction,’ which was compared to the channel layout of the array.

**Supplementary Figure 2.**
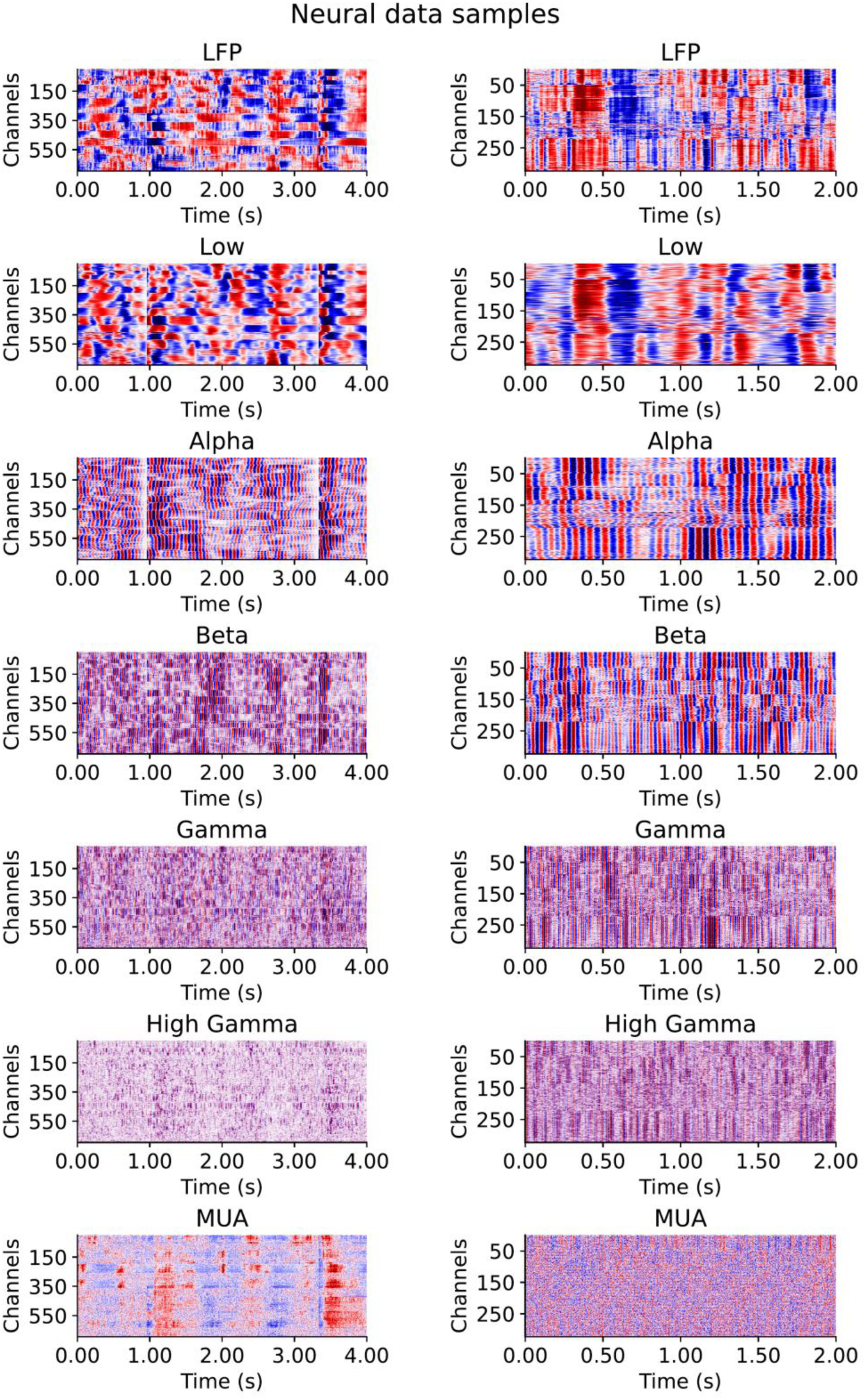
Eyes-closed LFP data filtered in different frequency bands, and MUA, across all recording sites, for monkeys L (left) and A (right).

**Supplementary Figure 3.**
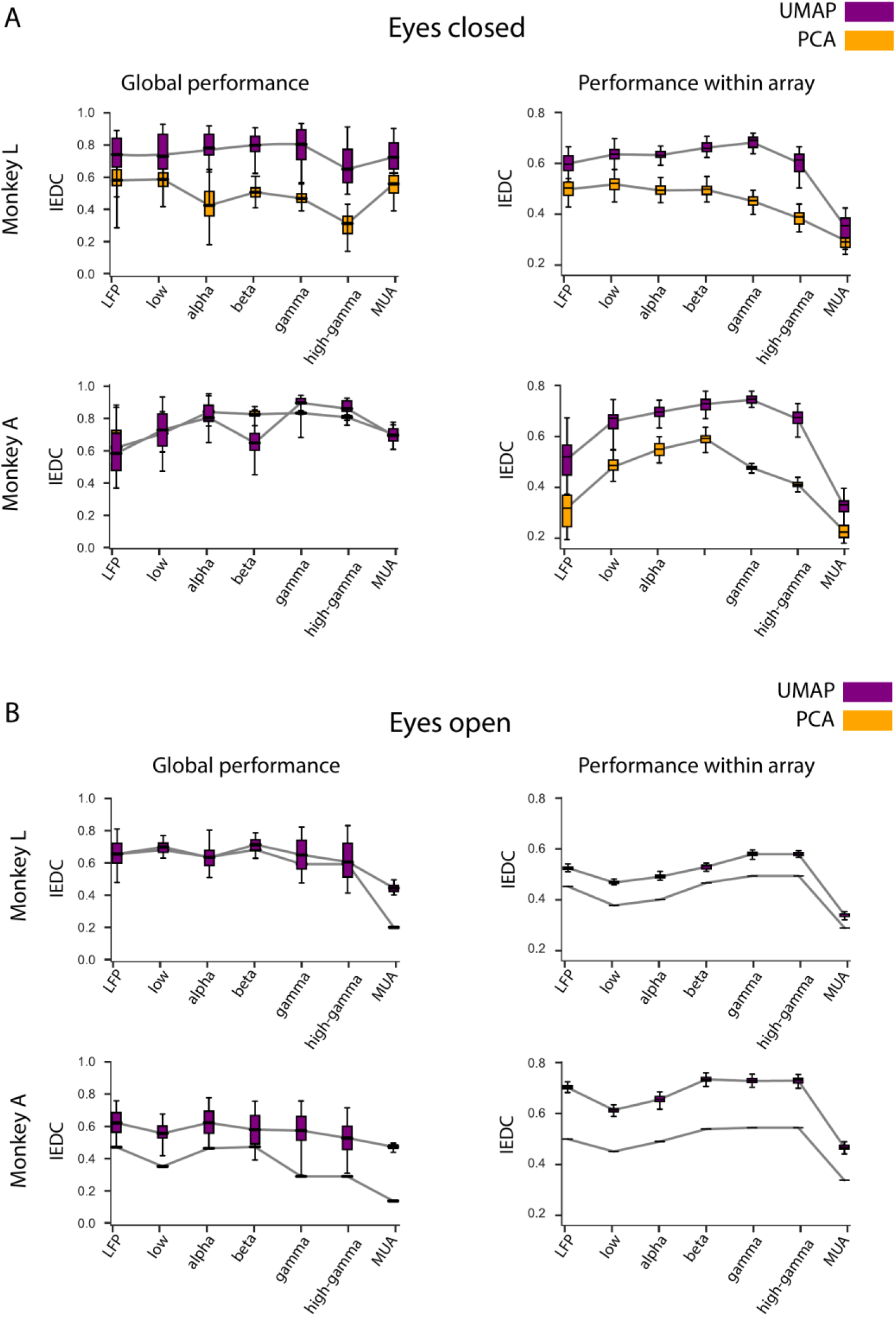
Performance of UMAP and PCA predictions (mean ± s.d.) using different types of data under eyes-closed (A) and eyes-open (B) conditions. IEDC values were generally higher with UMAP than with PCA.

**Supplementary Figure 4.**
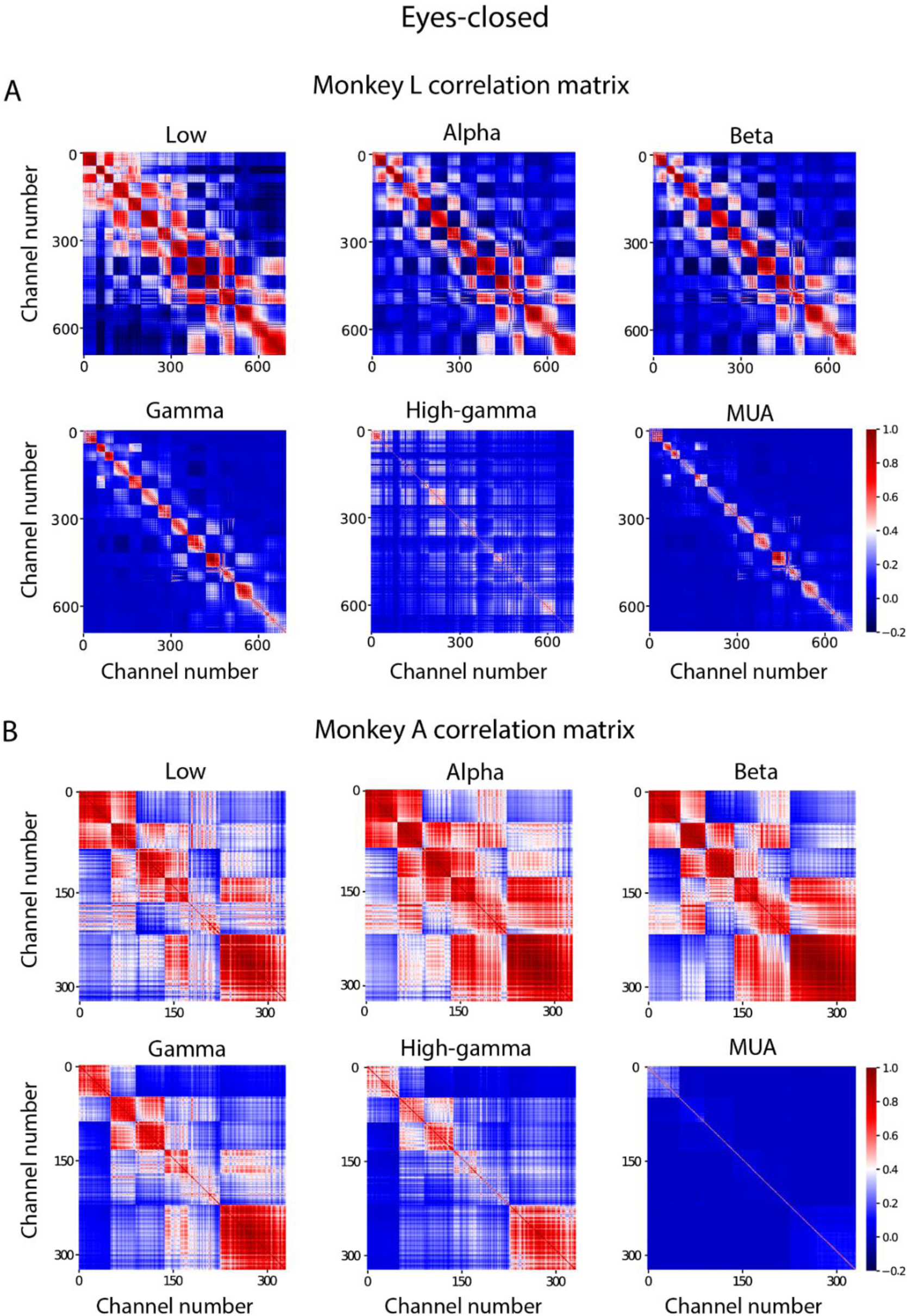
Correlation matrices for eyes-closed data in Monkeys L (A) and A (B): The colour of each entry i,j in the table represents the correlation between channels *i* and *j*.

**Supplementary Figure 5.**
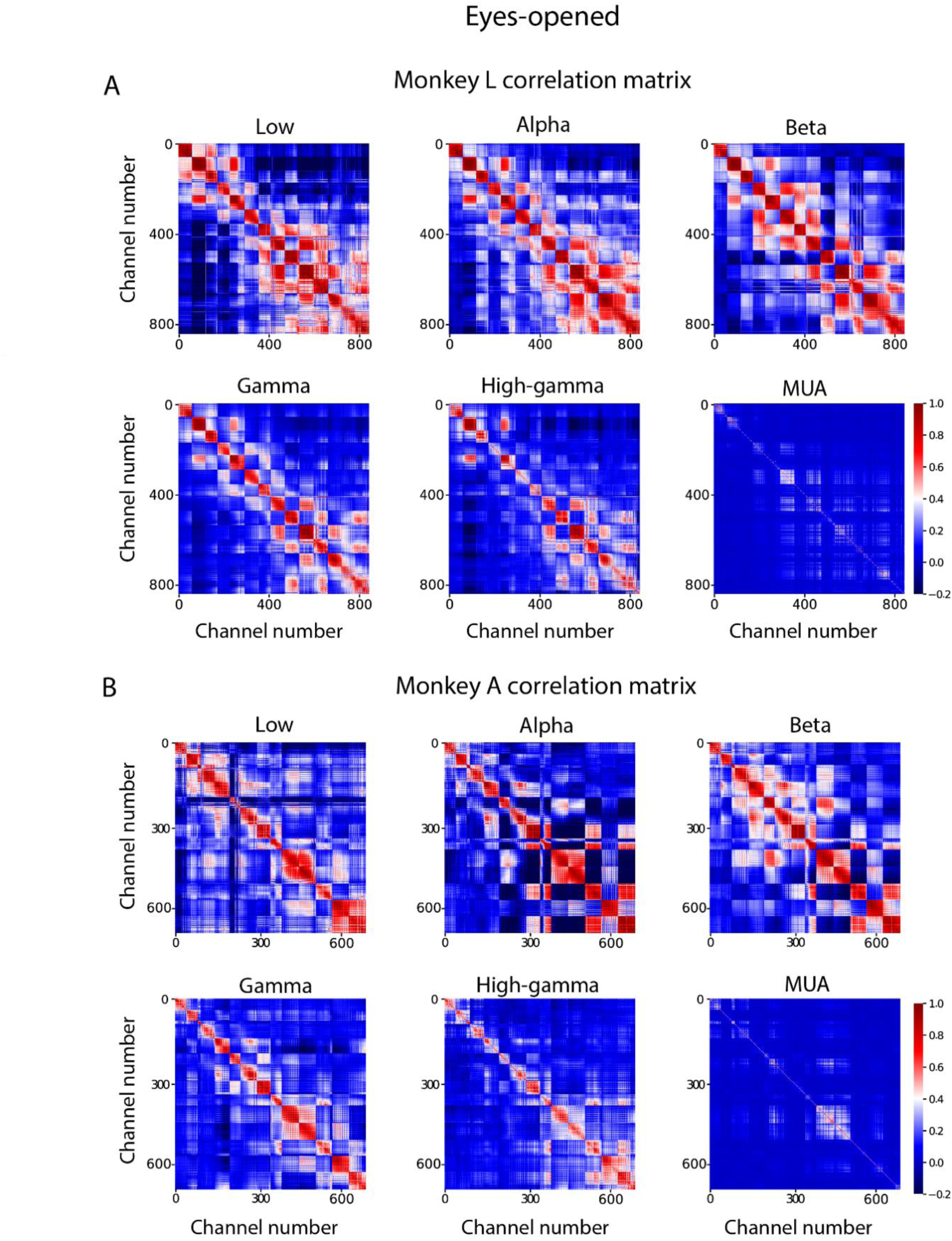
Correlation matrices for eyes-open data in Monkeys L (A) and A (B): The colour of each entry i,j in the table represents the correlation between channels *i* and *j*.

**Supplementary Figure 6.**
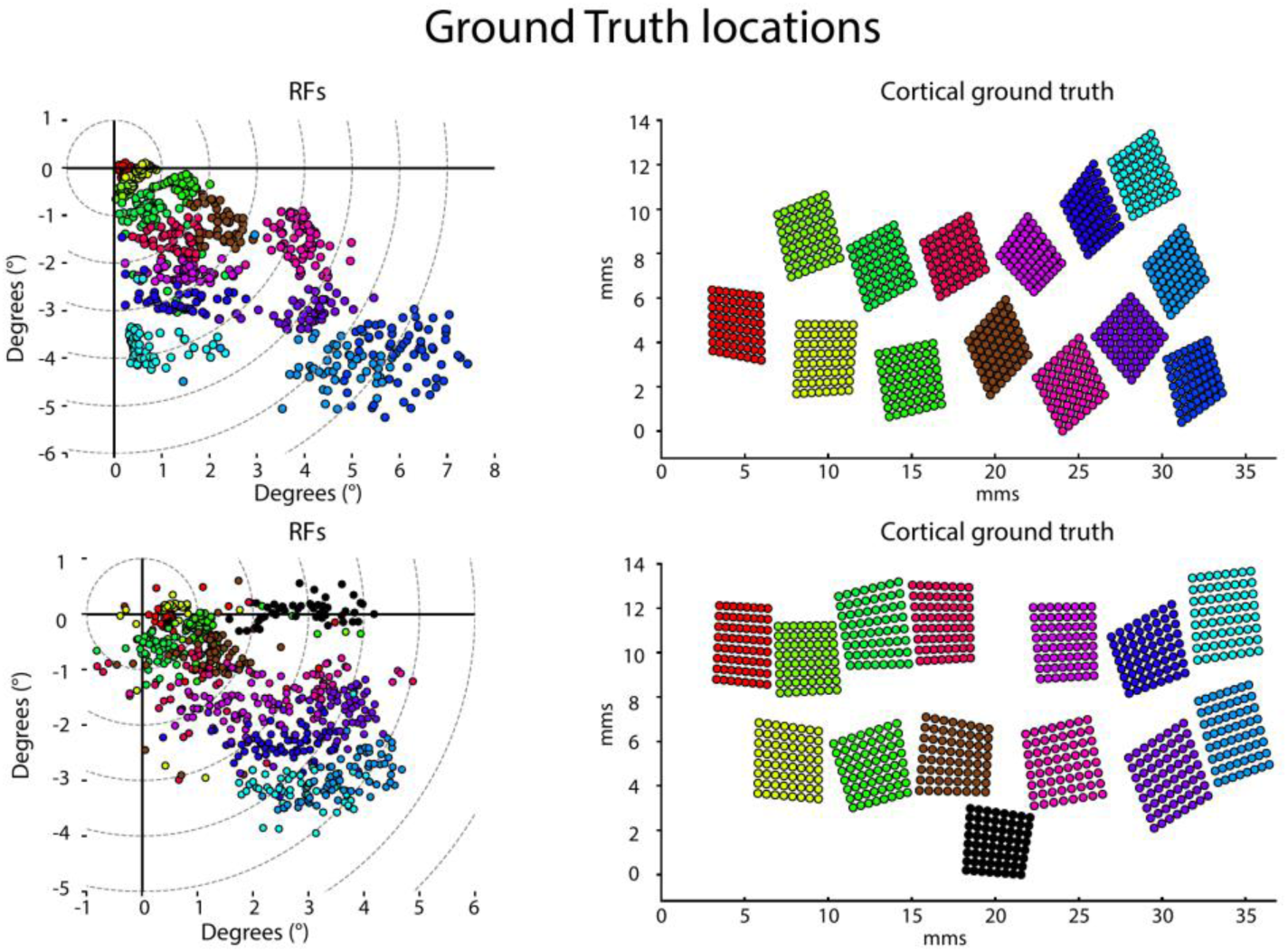
Ground truth locations of RF centres in visual space (left), and of electrodes and arrays in cortical space (right), for monkeys L and A. RFs are colour-coded according to the array to which they belong.

**Supplementary Figure 7.**
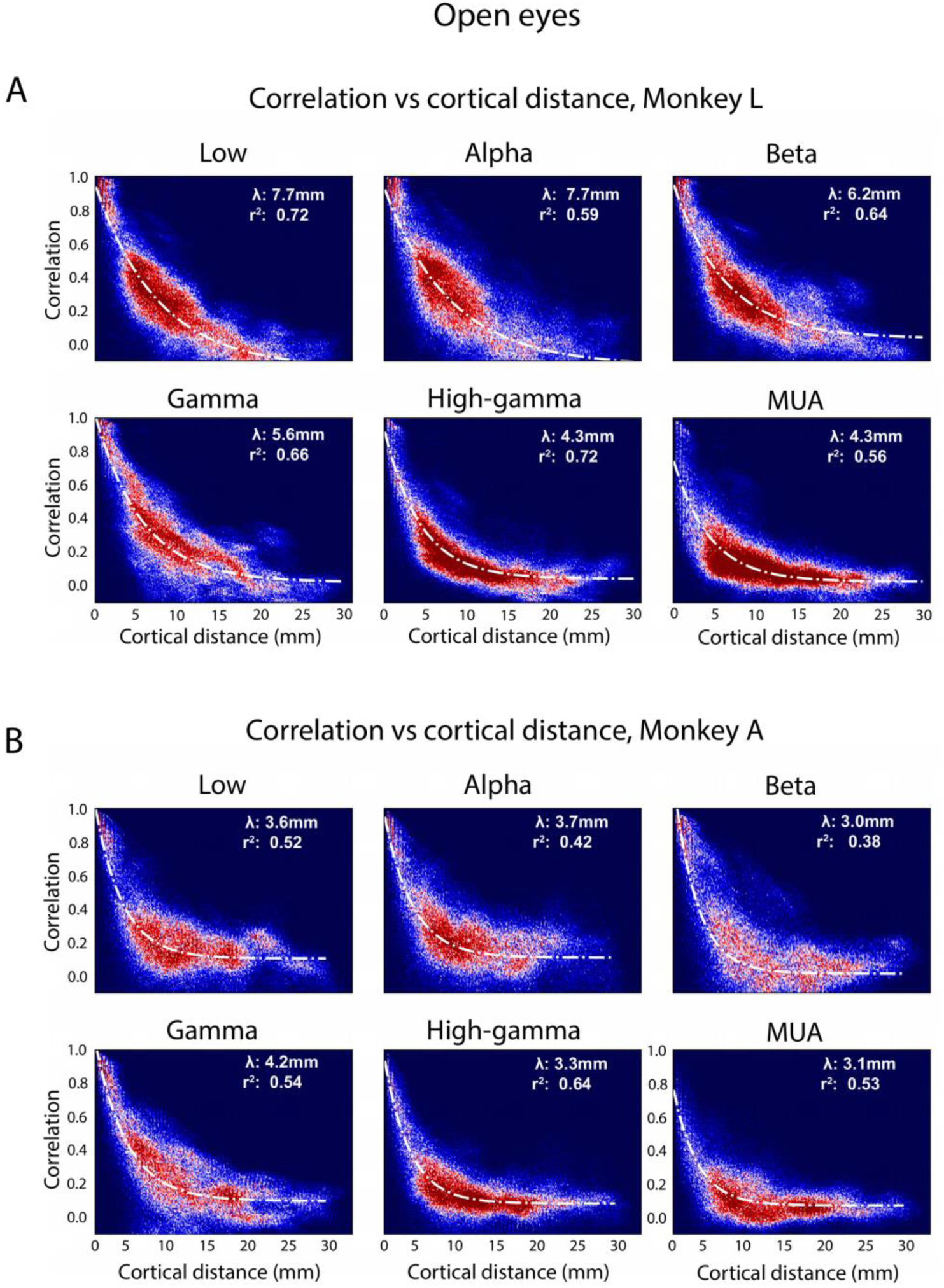
The relationship between pair-wise signal correlations and cortical distance for eyes-open data, for the different data-types in monkey L (A) and A (B). The fitted curve is an exponential.

**Supplementary Figure 8.**
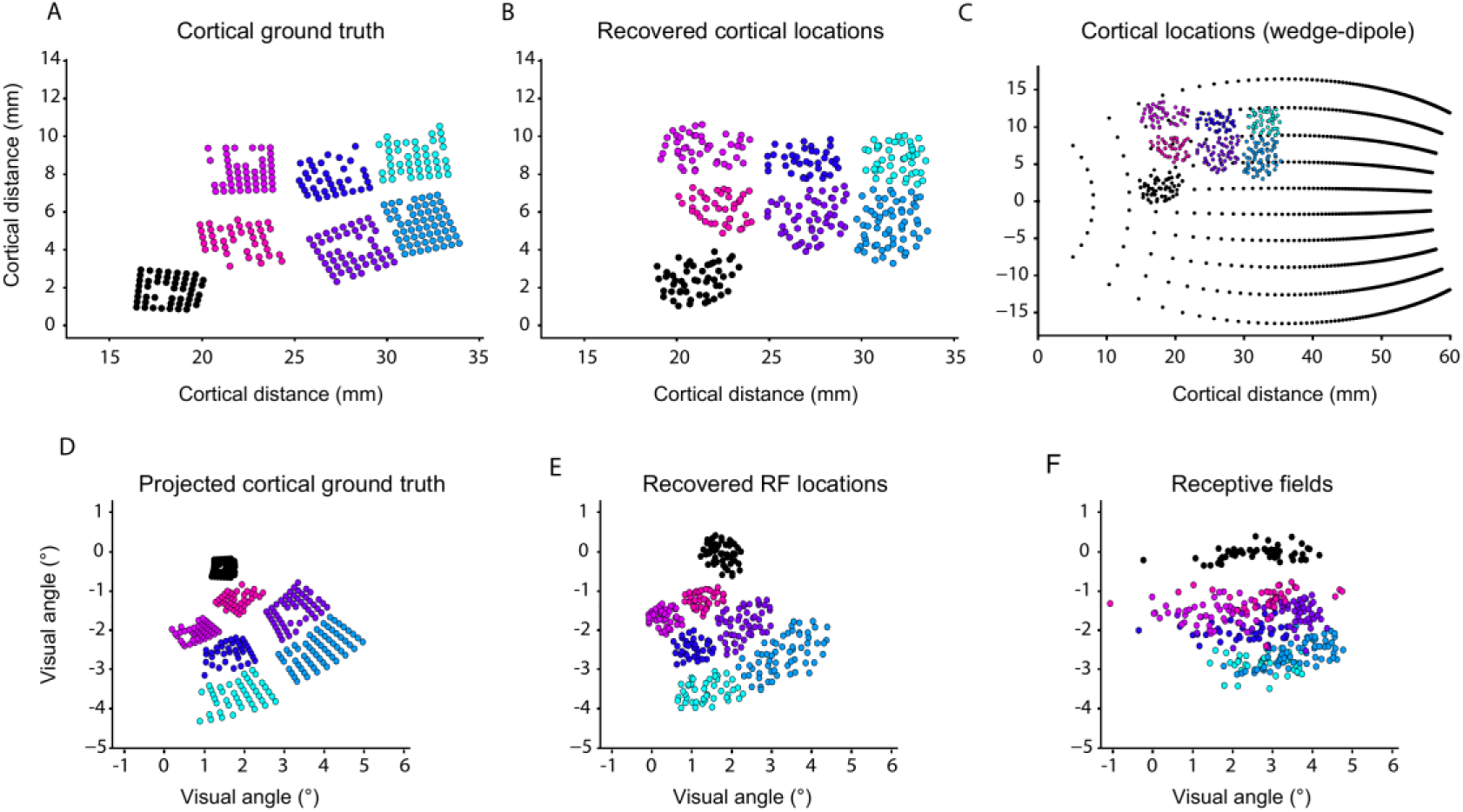
Transformation into retinal coordinates, monkey L, eyes-closed. A, Ground-truth positions of electrodes in the cortex in monkey L. B, NEUmap prediction in cortical coordinates. C, A wedge-dipole model was used to convert cortical coordinates into retinotopic space. D, Conversion of the cortical ground-truth map into retinal coordinates. E, Conversion of the NEUmap prediction into retinal coordinates. F, RFs measured with a moving bar stimulus while the monkey looked at a fixation spot.

**Supplementary Figure 9.**
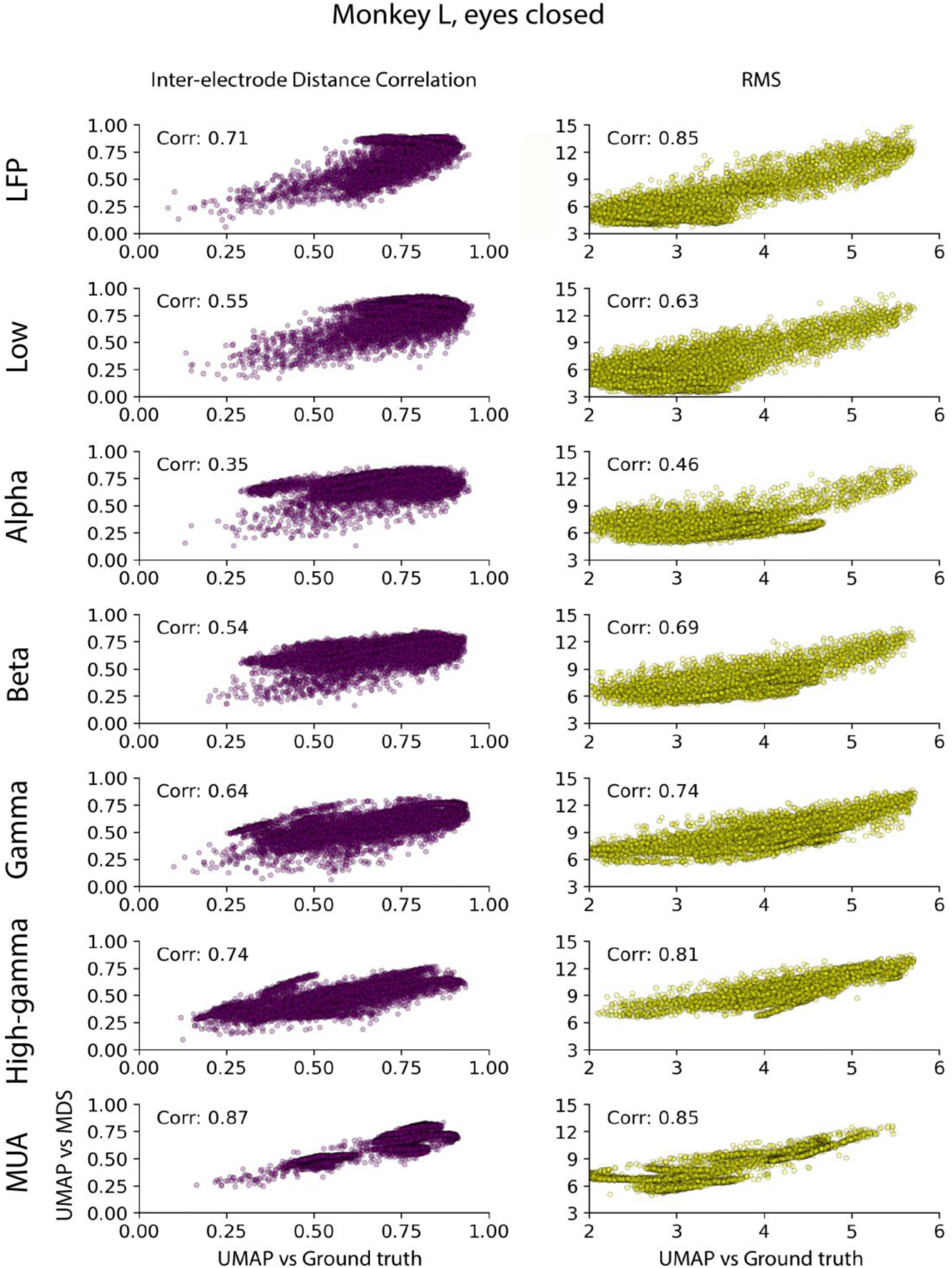
Correlations between performance metrics (left: IEDC, right, RMSE) comparing UMAP predictions and the ground truth cortical data (X-axis), and between UMAP predictions and Map_Prelim_ (Y-axis) based on MDS. Rows show different data-types; broadband LFP, low, alpha, beta, gamma, and high gamma frequencies and MUA.

**Supplementary Table 1.**
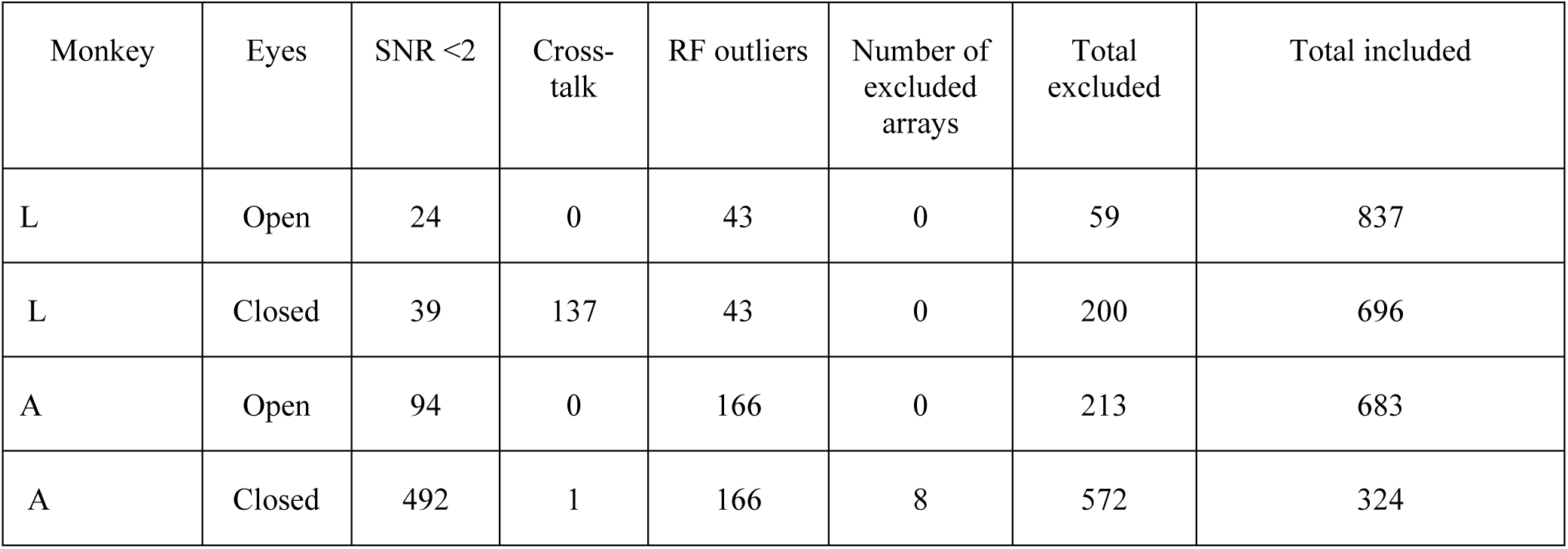
Numbers of excluded channels. Channels were excluded if they had an SNR < 2, if there was cross-talk or poorly defined RFs. We also excluded arrays if fewer than 25 channels with a sufficient SNR remained.

| Monkey | Eyes | SNR <2 | Cross-talk | RF outliers | Number of excluded arrays | Total excluded | Total included |
| --- | --- | --- | --- | --- | --- | --- | --- |
| L | Open | 24 | 0 | 43 | 0 | 59 | 837 |
| L | Closed | 39 | 137 | 43 | 0 | 200 | 696 |
| A | Open | 94 | 0 | 166 | 0 | 213 | 683 |
| A | Closed | 492 | 1 | 166 | 8 | 572 | 324 |

